# Transcranial Direct Current Stimulation to Modulate fMRI Drug Cue Reactivity in Methamphetamine Users: A Randomized Clinical Trial

**DOI:** 10.1101/2021.04.12.21255366

**Authors:** Hamed Ekhtiari, Ghazaleh Soleimani, Rayus Kuplicki, Hung-Wen Yeh, Yoon-Hee Cha, Martin Paulus

## Abstract

Transcranial direct current stimulation (tDCS) has been studied as an adjunctive therapeutic option to alter maladaptive cortical excitability, activity, and connectivity associated with chronic substance use via the application of a weak direct current through the brain. The underlying mechanism of action remains ambiguous, however. We present a randomized, triple-blind, sham-controlled, clinical trial with two parallel arms conducted to determine the neural substrates of tDCS effects on drug craving using an fMRI drug cue reactivity paradigm. Sixty participants with methamphetamine use disorder were randomly assigned to two groups: 30 participants to active tDCS (5×7 cm^2^, 2 mA, for 20 minutes, anode/cathode over the F4/Fp1 in EEG 10-20 standard system) and 30 participants to the sham group. Neuroimaging data of a methamphetamine cue reactivity (MCR) task were collected immediately before and after stimulation with subjective craving assessed before, after, and during fMRI scans. There was a significant reduction in self-reported craving after stimulation (main effect of time) without any significant effect of group, time, or by group-time interaction. Our whole-brain analysis demonstrated that brain activation decreased in all parts of the brain in the second (post-stimulation) MCR imaging session after sham stimulation (habituation) but this uniform decrease did not occur throughout the brain in the active group. There were significant interactions between the group (active vs. sham) and time (after vs. before stimulation) in five main regions; medial frontal gyrus, anterior insula, inferior parietal lobule, precuneus, and inferior frontal gyrus with higher activations after active stimulation. We simulated computational head models for each individual. There was a significant effect of group in the relationship between level of current in the above-mentioned significant clusters and changes in task-modulated activation. We also found that brain regions with the highest electric fields in the prefrontal cortex showed a significant time by group interaction in task-modulated connectivity (psychophysiological interaction during MCR) in the frontoparietal network. In this two-parallel-arms triple-blind randomized control trial, we did not find any significant effect of the one session of active F4/Fp1 tDCS on drug craving self-report compared to sham stimulation. However, connectivity differences induced by active compared to sham stimulation suggested some potential mechanisms of tDCS to modulate neural response to drug cues among people with methamphetamine use disorder.

**Highlights:**

- No significant effect of active stimulation compared to sham was found in self-reported craving.
- Uniform habituation in response to drug cues happens only after the sham stimulation.
- MFG, IFG, insula, IPL, and precuneus show significantly higher responses to cues after active stimulation.
- Head models showed our stimulation montage (F4-Fp1) induces the highest level of current in rSFG.
- Cue reactivity modulated connectivity was significantly reduced by active stimulation between rSFG and rPPC.

## Introduction

Amphetamines, including methamphetamine, constitute the second most commonly used group of illicit substances worldwide, with 13.9 million to 54.8 million estimated users (United Nations Office on Drugs and Crime, 2014; http://www.unodc.org) facing significant morbidity and mortality (Han et al. 2021). Methamphetamine activates intra and extracellular pathways that induce both neurotoxic and neuroadaptive changes in the brain (Yang et al. 2018, Shaerzadeh et al. 2018, Moratalla, Ares-Santos, and Granado 2014). There is limited reliable evidence from randomized double-blind controlled clinical trials for the effectiveness of pharmacological or non-pharmacological interventions in methamphetamine use disorders (MUDs)(Trivedi et al. 2021). Therefore, there is an urgent need to develop new therapeutic interventions for MUDs. Recent advancements in human neuroscience research have provided an emerging line of non-invasive brain stimulation interventions for targeting the neurocognitive processes underlying substance use disorder (SUD) in general and MUD in particular, the effectiveness of which is still under investigation (Verdejo-Garcia et al. 2019, Ekhtiari et al. 2019a).

Drug craving has been considered to be one of the core processes that contributes to drug seeking behavior and relapse when testing the efficacy of interventions for SUDs. Drug cue exposure is well-known as an ecologically valid paradigm to induce drug craving in the experimental setting (Ekhtiari et al. 2016). Previous fMRI studies reported that cue exposure in people with SUDs is associated with alterations in brain functions that effectively engage different large-scale brain networks related to the reward, habit, salience, executive, memory, and self-directed processing during drug cue exposure (Zilverstand et al. 2018). In this context, reduction of cue induced craving during a cue exposure task and modulation of its neural processing circuits is considered to be a critical target for enhancing substance use recovery (Courtney, Ghahremani, and Ray 2016, Courtney et al. 2016).

Modulation of prefrontal cortical areas that act as main hubs in the neural processing of exposure to drug cues and subsequent craving is a primary strategy of noninvasive brain stimulation studies for SUDs including those that use transcranial direct current (tDCS) and transcranial magnetic stimulation (TMS). There is preliminary but promising evidence that both tDCS and TMS can be effective in modulating drug craving (Jansen et al. 2013). However, the evidence is very limited in MUD. A recent systematic review in 2019 reported that only 6 out of 50 TMS studies and 3 out of 34 tDCS studies in the field of SUD were performed in participants with MUDs (Ekhtiari et al. 2019b).

tDCS is an easy-to-use technology that is becoming increasingly popular because of its low cost and high availability. One of the first sham-controlled crossover tDCS studies included thirty abstinent male methamphetamine users and investigated the immediate online state-dependent effect of tDCS on cue-induced methamphetamine craving using a computerized cue induced craving task (Shahbabaie et al. 2014, Alireza Shahbabaie 2014). That study demonstrated that when compared to sham, active tDCS led to a larger decrease of self-reported craving at rest and induced larger craving ratings during cue exposure. Recent research combined tDCS with computerized cognitive addiction therapy (CCAT) to study the synergistic effects of bilateral tDCS along with cognitive training in 73 female chronic methamphetamine users using a parallel design trial with 24 subjects in CCAT + tDCS group, 26 subjects in CCAT + sham tDCS group, and 23 subjects in control group. After 20 sessions of CCAT + tDCS (active or sham) intervention a significant reduction in craving in the active group compared to sham was reported. However, to the best of our knowledge, there is only one study in MUDs that combined tDCS with neuroimaging to study the neural effect of tDCS in MUDs (Shahbabaie et al. 2018). In this study, fMRI was done immediately before and after stimulation in a crossover, double-blind, sham-controlled trial in fifteen male participants with MUDs. The authors reported that subjective craving decreased significantly after active tDCS (bifrontal, F4-F3) compared to sham tDCS while showing that resting-state functional networks including default mode, executive control, and salience networks were significantly modulated by active tDCS. Additionally, alteration of self-reported craving score after stimulation was significantly correlated with the modulation of these large-scale resting-state networks. In a secondary exploratory analysis among 10 participants, the authors demonstrated a significant correlation between changes in functional activity and individualized electric field within the frontopolar cortex (Esmaeilpour et al. 2019). These preliminary findings motivated our larger scale study with pre-defined hypotheses for both activation and connectivity modulation.

To better understand the mechanisms involved in tDCS modulated drug craving, we studied the effects of tDCS on drug cue reactivity and craving using fMRI immediately before and after stimulation in a randomized, triple-blind sham-controlled trial of 60 participants with MUDs during the early abstinence phase of their participation in residential recovery program. In a parallel design, we applied unilateral right DLPFC stimulation (F4) or a matched sham stimulation protocol with the return electrode on the contralateral supraorbital area (FP1). We have simulated individualized computational head models to integrate the personalized regional electrical current dose in the prediction of response to tDCS. In fMRI analysis as well as subjective reports, we tested whether tDCS over DLPFC would modulate cue induced craving, hypothesizing that such an effect would be larger in the active group with brain regions that had the highest concentration of current density playing a crucial role in response to cue exposure. To the best of our knowledge, this is the first triple-blind tDCS-fMRI clinical trial in the field of MUDs that contains a relatively large (60 participants) sample with self-report and fMRI data. These data were used to assess tDCS effects on activation and functional connectivity during cue exposure along with computational head modeling to quantify the relationship between the estimated induced electric field (EF) and drug cue reactivity outcomes.

## Method

### Participants

A total of 80 potential participants were screened for eligibility in this trial. Four individuals were excluded for fMRI safety criteria and one individual was excluded for not being able to follow instructions (cognitive deficits). Seventy-five individuals were consented. Eight individuals were lost in follow up before randomization and seven individuals were removed due to technical problems in the scanning or stimulation sessions. Sixty participants (all-male, mean age ± standard deviation (SD) = 35.86 ± 8.47 years range from 20 to 55) with methamphetamine use disorder (MUD) were included in the final randomization phase. All completed imaging and stimulation sessions and contributed data for analysis. All participants were recruited (from October 2017 to January 2019) during their early abstinence period from the 12&12 residential drug addiction treatment center in Tulsa, Oklahoma. The trial protocol, primary outcome (change in craving self-report and cue induced brain activation), and secondary outcomes were preregistered in ClinicalTrials.gov (Identifier: NCT03382379). Written informed consent was obtained from all participants before participation and the study was approved by the Western IRB (WIRB Protocol #20171742). This study was conducted in accordance with the Declaration of Helsinki and all methods were carried out in accordance with relevant guidelines and regulations.

The inclusion criteria for this study were: (1) English speaking, (2) diagnosed with methamphetamine use disorder in the last 12 months, (3) admitted to a residential abstinence-based treatment program for methamphetamine use disorder, (4) abstinence from methamphetamine for at least one week, and (5) willing and capable of interacting with the informed consent process. Exclusion criteria included: (1) unwillingness or inability to complete any of the major aspects of the study protocol, including magnetic resonance imaging (i.e. due to claustrophobia), drug cue rating or behavioral assessment, (2) abstinence from methamphetamine for more than 6 months based on self-report, (3) schizophrenia or bipolar disorder based on the MINI interview, (4) active suicidal ideation with intent or plan determined by self-report or assessment by the principal investigator or study staff during the initial screening or any other phase of the study, (5) positive drug test for amphetamines, opioids, cannabis, alcohol, phencyclidine, or cocaine confirmed by breath analyzer and urine tests.

### Data acquisition procedure

The data acquisition procedure is illustrated in *Figure 1*. This study was a randomized, triple-blind, sham-controlled, clinical trial with two parallel arms. After obtaining their written consent, each participant was randomized during the baseline session to receiving active or sham stimulation using a computer-generated program that allocated participants into blocks of four. This protocol assured that both participants and data collectors were kept blind to stimulation conditions. A coding system developed by the tDCS device manufacturing company (NeuroConn®) supported blinding during data collection for both the research team and participants. After completion of data collection, a list of participants’ codes for active and sham stimulation was prepared. A separate person who did not participate in data collection or analysis had access to the randomization information and prepared a set of non-identifying codes (A and B) for analysis. Unblinding occurred after completion of data analysis and preparation of the initial draft of the results’ figures and tables. The preprocessed sample size of 30 participants per arm provided 80% power to detect an effect size (Cohen’s d) of 0.74 for changes in drug-cue reactivity between each arm at a two-sided 0.05 significance level in a two-sample t-test.

**Figure 1:**
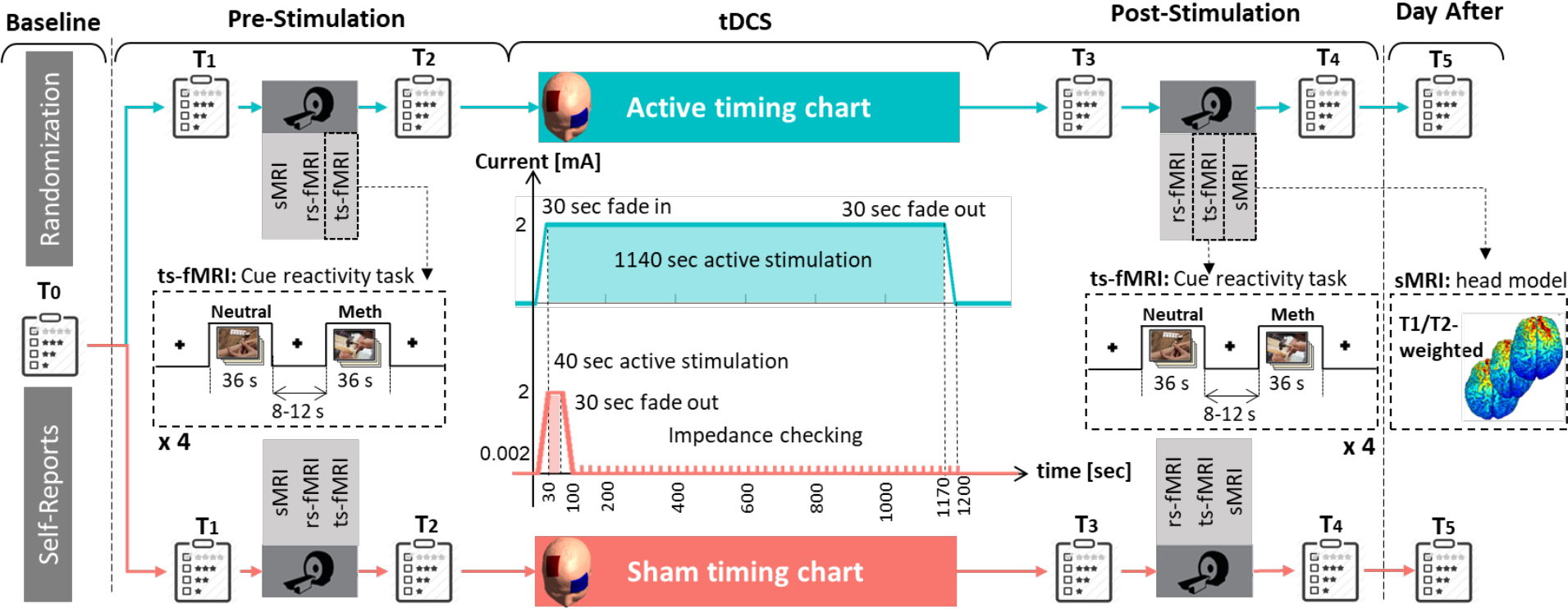
Data acquisition procedure. At baseline (before stimulation session), 60 participants with methamphetamine use disorder were randomly assigned to active or sham transcranial direct current stimulation (tDCS): 30 participants in each group. In this parallel design triple-blind sham-controlled trial, neuroimaging data including structural MRI (sMRI), resting-state (rs-fMRI), and task-based (ts-fMRI) fMRI data were collected in a pre-stimulation/post-stimulation design. fMRI task was a standard cue-reactivity task with a pictorial block design that consisted of methamphetamine vs neutral stimuli and sMRI data included T1- and T2-weighted images that were used for creating individualized computational head models (CHMs). Self-report data were also collected at 6 time points: baseline (T0), immediately before (T1, T3) and after (T2, T4) each MRI acquisition, and the day after stimulation (T5). Stimulation at 2 mA intensity was delivered through 5×7 cm^2^ sponge electrodes with the anode over F4 and cathode over Fp1. Neuroconn stimulator was used for both active (in blue) and sham (in red) stimulation and timing charts are represented in the center of the figure. Fade in and fade out were 30 sec in both groups. Participants in the active stimulation group received 2 mA stimulation during 1140 sec whereas participants in the sham group received 40 sec of 2 mA stimulation. Fade out in the sham group was followed by 1100 sec without any stimulation (impedance was controlled so that average current overtime was not more than 2 *μ*A). Abbreviation: sMRI: structural MRI, rs-fMRI: resting-state fMRI, ts-fMRI: task-based fMRI, CHM: computational head model.

Demographic and substance use profiles of each group are presented in *Table 1*. Neuroimaging data including structural MRI (T1- and T2-weighted MRI), resting-state, and task-based (a block design cue-reactivity task (Ekhtiari, Kuplicki, Aupperle, et al. 2020)) fMRI data were collected in a pre-stimulation/post-stimulation design. Subjective craving for methamphetamine was assessed by Visual Analogue Scale (VAS) (scored 0-100) at 6 time points; baseline (T0), immediately before (T1 and T3) and after (T2 and T4) each fMRI scan, and the day after (T5) the data collection. Adverse effects of tDCS were monitored by asking participants after active or sham stimulation whether they had experienced any side effects with a questionnaire containing a Likert scale from 0 to 5 (0 = none, 1 = very mildly, 2 = mildly, 3= moderate, 4 = severe, 5 = very severe) for 10 symptoms at the end of the scanning session and on the next day. Additionally, to check the efficiency of the blinding, at the end of the stimulation session participants were asked about whether they believed they received sham or active stimulation and how confident they were in that assessment.

**Table 1:** Demographic data. Between-group (Active/Sham) differences are reported based on uncorrected p-values.

| | Mean (SD) | | $p^{a,b}$<br>value |
| --- | --- | --- | --- |
|  | Sham group | Active group |  |
| Age | 37.75 (8.78) | 34.96 (7.92) | 0.202 |
| Education | 13.17 (2.94) | 13.73 (2.49) | 0.963 |
| BMI | 28.16 (4.12) | 26.98 (5.78) | 0.710 |
| Age of Meth use onset | 20.18 (6.72) | 20.58 (7.71) | 0.834 |
| Duration of Meth use at least once a week (years) | 16.44 (27.37) | 9.67 (8.92) | 0.203 |
| Cost of Meth (dollar per month) | 1250.8 (1448) | 986.17 (1146) | 0.436 |
| Dose of Meth (gram per day) | 1.47 (1.30) | 1.74 (1.83) | 0.498 |
| History of Meth injection (n (%)) | 23 (76.7%) | 23 (76.7%) | 1.000 |
| **Days of Drug use in the last month (before abstinence)** |  |  |  |
| Meth | 25.33 (7.9) | 21.27 (10.30) | 0.071 |
| Alcohol | 10.27 (11.30) | 8.47 (12.03) | 0.552 |
| Heroin | 4.27 (10.00) | 4.33 (9.04) | 0.978 |
| Methadone | 0.1 (0.55) | 1.03 (5.47) | 0.357 |
| Other opioids | 2.57 (6.38) | 2.30 (5.75) | 0.866 |
| Barbiturate | 0.17 (0.91) | 0.00 (0.00) | 0.326 |
| Sedative | 0.67 (1.6) | 4.03 (8.81) | 0.048 |
| Cocaine | 0.3 (1.02) | 0.17 (0.75) | 0.566 |
| Cannabis | 9.53 (11.33) | 6.07 (8.37) | 0.183 |
| Hallucinogens | 0.07 (0.25) | 0.7 (2.85) | 0.236 |
| Inhalants | 0.07 (0.37) | 0.03 (0.18) | 0.656 |
| Duration of current abstinence (days) | 58.33 (30.21) | 63.27 (39.56) | 0.556 |
| Meth Cue Reactivity Screening score (MCRS) score (0-100) | 58.82 (25.41) | 63.28 (20.16) | 0.313 |
| Meth Withdrawal Scale score (0-24) | 2.67 (3.04) | 4.67 (5.76) | 0.100 |

<sup>a</sup>: Independent sample t-test, <sup>b</sup>: Chi-square test

In order to reduce variability across the population due to impact of time of the day on stimulation effects and outcomes, all 60 participants received stimulation (active or sham) over the DLPFC in the afternoon (from 3:00 pm to 7:30 pm). The time of stimulation was recorded for each individual. The distribution of stimulation timing can be found in *Figure S1*.

### Transcranial direct current stimulation

tDCS was applied via two saline-soaked surface sponge electrodes (area = 5×7 cm^2^) that were connected to the battery-driven NeuroConn DC stimulator MR. In order to target the right DLPFC unilaterally, the “Beam F3-system” (Rossini et al. 2015) was used based on nasion-to-inion and tragus-to-tragus distance, and head circumference in order to calculate electrode coordinates over the scalp for each individual. The anode was placed over F4 (in EEG 10-20 standard system) with the long axis of the pad pointing towards the vertex of the head. The cathode electrode was positioned over the contralateral eyebrow (Fp1 EEG electrode site also referred to as the supraorbital position) with the long axis of the pad parallel to the horizontal plane. The electrodes were fixed to the scalp using multiple rubber head bands. In order to recheck the orientation as well as the location of the electrodes, and between-electrode distance, one photo was taken without showing the participants’ face or any identifiable information (e.g., scars or tattoos). This photo was rechecked by an expert to exclude any potential individuals with inaccurate electrode placement.

Timing charts of active and sham stimulation are illustrated in the center of *Figure 1.* For active stimulation, tDCS was delivered for 20 minutes at an intensity of 2 mA with 30 sec ramp-like fade-in, 1140 sec active stimulation, and 30-sec ramp-like fade-out. For the sham stimulation procedure, the stimulator automatically switched off after 100 sec (30-sec ramp-like fade-in, 40-sec active stimulation, and 30-sec ramp-like fade-out) yielding sensations typically elicited by tDCS. Fade-out in the sham group was followed by 1100 sec without any stimulation (just impedance was checked periodically and the average current over time was not more than 2 μA). Impedance was kept below 10 k-Ohm during both active and sham tDCS.

### Structural MRI data acquisition parameters

As shown in *Figure 1*, MR images were collected immediately before and after stimulation. Structural and functional MRIs were obtained on two identical GE MRI 750 3T scanners. High resolution structural images were acquired through magnetization-prepared rapid acquisition with gradient-echo (MPRAGE) sequence using the following parameters: TR/TE = 5/2.012 ms, FOV/slice = 24 x 192/0.9 mm, 256×256 matrix producing 0.938 x 0.9 mm voxels and 186 axial slices for T1-weighted images and TR/TE=8108/137.728ms, FOV/slice=240/2mm, 512×512 matrix producing 0.469×0.469×2mm voxels and 80 coronal slices for T2-weighted images. T1- and T2-weighted MR images were used for generating computational head models (CHMs) for each individual.

### fMRI data collection procedure

Resting-state fMRI data were acquired with one 8-minute run (TR/TE = 2000/27 ms, FOV/slice = 240/2.9 mm, 128×128 matrix producing 1.857×1.857×2.9 mm voxels, 39 axial slices, and 240 repetitions) pre- and post-stimulation with instructions to the participant to keep their eyes fixated on a cross presented on the screen and to try not to think of anything in particular.

The methamphetamine cue reactivity (MCR) task was administered after each resting state scan and had the same parameters, except containing 196 repetitions. The MCR task contained pictorial methamphetamine cue exposures in a block design with two sets (methamphetamine and neutral) of distinct but equivalent pictures validated in another study (Ekhtiari, Kuplicki, Pruthi, et al. 2020). The total task time was approximately 6.5 minutes and contained 4 neutral and 4 meth picture blocks. Each block included a series of 6 pictures of the same category (meth or neutral) each was presented for 5 seconds with a 0.2 second blank inter-stimulus interval. A visual fixation point was presented for 8 to 12 seconds between each block. A meth craving inquiry followed each block (meth or neutral) in which participants were asked to rate their current meth craving level on a 1 to 4 rating scale (1 lowest to 4 highest craving level). Participants’ response times to the rating in each block were recorded. Furthermore, within each block, a yellow box was presented around one of the six pictures. Participants were asked to press a button as soon as they saw the box; their reaction time was recorded. The task paradigm is illustrated in *Figure S2* with a detailed explanation.

### Task-based functional activity analysis

Functional data analysis was performed in AFNI. The first three pre-steady state images were removed. The preprocessing steps were as follows: despiking, slice timing correction, realignment, transformation to MNI space, and 4 mm of Gaussian FWHM smoothing. Three polynomial terms and the six motion parameters were regressed out. TRs with excessive motion (defined as the Euclidian norm of the derivative of the six motion parameters being greater than 0.3) were censored during regression.

A linear mixed-effect model (LME) using 3dLME in AFNI was used to study the effects of tDCS on the whole brain functional activation. As fixed effects, we included the effects of time (pre/post-stimulation), group (real/sham), and their interaction. A random intercept was included for each subject. Post- and pre-stimulation functional activity were compared by using LMEs (3dLME, AFNI). Family-wise error (FWE) was found by Monte-Carlo simulation-based (3dClustSim, AFNI) multiple comparison correction with alpha > 0.1. P < 0.005 and cluster size > 40 was considered for reporting the results.

In order to search for the overall stimulation effects on the direction of changes in brain functional activity, brain activation during the drug cue reactivity task was calculated for meth > neutral contrast before and after stimulation across the population. For each participant, mean beta weight values were estimated for all ROIs in the Brainnetome atlas (BNA) (Fan et al. 2016). Change in the ROI-wise activation over time was analyzed with an LME using “nlme” package for linear mixed modeling in R software (v.1.2.5) and included using fixed effects of time, group, and interaction between group and time. By-subject intercept and ROI terms (246 ROIs in BNA) were entered as random effects.

### Task-modulated functional connectivity analysis

For task-modulated connectivity analysis, preprocessing of the MRI data including structural, and task-based fMRI data was performed using the CONN toolbox (v.20.b) (Whitfield-Gabrieli and Nieto-Castanon 2012). Images were reoriented to the anterior commissure and co-registered to the T1 image. Segmentation and normalization of the T1-weighted images were performed using CONN and each normalized image was checked to detect incoherent deformations.

Task-based data underwent a standard preprocessing pipeline in CONN which included slice timing correction, outlier identification to remove artifacts from fMRI data using Artifact Detection Tool (ART— an SPM package implemented in the CONN pipeline to remove signal intensity spikes and fMRI volumes with the excessive motion from the scan), segmentation and normalization into MNI standard space, and functional smoothing with an 8mm full width at half maximum (FWHM) Gaussian kernel.

To investigate task-modulated changes in connectivity, a seed to whole-brain analysis was performed to calculate generalized psychophysiological interaction (gPPI during the task). PPI is a task modulated connectivity calculation which identifies voxels in the brain that alter their connectivity with a seed region of interest in a given context (here the drug cue reactivity task). In the first step, a seed region was defined for gPPI analysis using electric field (EF) distribution patterns obtained from the group-level analysis of computational head models (CHMs). With respect to the blinding during the analysis step, individualized head models were generated for all 60 participants. CHMs were transformed to the standard fsaverage space for group-level analysis. Cortical atlas parcellation was used to calculate EFs in each brain sub-region in BNA. Averaged EFs, as an indicator of possible neuromodulation intensity, was calculated in all cortical sub-regions across the population. The brain region with the highest averaged EF was selected as a seed for the seed to whole-brain gPPI analysis. In the next step, BOLD signals were extracted from the BNA seed region and seed-to-whole brain task-modulated connectivity was calculated.

In the first-level analysis, after defining the contrast of interest (meth > neutral), the gPPI design matrix including the seed region’s time course (physiological term), task time course (psychological term), the interaction between task and BOLD signal in the seed region (PPI term), and motion covariates was defined. For the second-level analysis, time (post vs pre) by group (active vs sham) interaction was calculated. Connection level threshold was p uncorrected < 0.001. Omnibus F test was used at the cluster level and p FDR corrected < 0.05 was considered as the threshold value for reporting results.

### Generating individualized computational head models

Gyri-precise CHMs were generated from a combination of high-resolution T1- and T2-weighted MR images for all participants. For a subset of participants (N = 8), T2-weighted MRIs were not available and head models were created only based on T1 images. As a part of the previous study, head models were generated for all of the participants to visualize how current flows through the brain using SimNIBS software. Briefly, automated tissue segmentation was performed in SPM12. The head volume was assigned to six major head tissues (white matter (WM), gray matter (GM), cerebrospinal fluid (CSF), skull, scalp, and eyeballs). The assigned isotropic conductivity values were WM = 0.126 Siemens/meter (S/m), GM = 0.275 S/m, CSF = 1.654 S/m, skull = 0.01 S/m, skin = 0.465 S/m, and eyeballs = 0.5 S/m. The results were visualized using Gmsh and MATLAB.

### Self-report data analysis

Craving score immediately before and after the MRI session was collected outside the scanner. In 6 time points (T0 to T5), participants reported their cravings for drug using a Visual Analog Scale (VAS). The desire for Drug Questionnaire (DDQ) (Franken, Hendriks, and van den Brink 2002) with three main sub-scores, i.e., desire and intention, negative, and control was collected in four different time points (T0, T1, T4, and T5). As described in the task design section, inside scanner craving scores, reaction time to the rating, and response time to a yellow box were also collected during the cue-induced craving task. LME models (fixed effect: time, group, and time by group interaction; random effect: subject) with post-hoc analysis was used to investigate changes in self-reports from before to after intervention. Furthermore, adverse effects of tDCS and efficacy of blindness were compared between the two groups. P uncorrected < 0.05 was considered as the threshold value in reporting behavioral data.

### Correlation/regression analysis

Based on functional activity analysis, we tested for a relationship between induced EFs and functional activity within the clusters with significant time by group interactions using regression models with changes in functional activity as the dependent variable. We also considered EFs within the cluster and group as two independent regressors. Additionally, in order to test the relationship between craving and functional activity, we calculated correlation between craving changes ((T2 – T1) as well as (T4 – T3)) and functional activity and EFs within the significant clusters.

With respect to the connectivity analysis, we used regression models with changes in functional connectivity as dependent variable and group and EFs as regressors. We also used correlation analysis to determine the relationship between craving changes and functional connectivity between the seed region and clusters that showed significant time by group interaction in terms of task modulated connectivity.

## Results

Demographic data collected at the baseline can be found in *Table 1* for all participants in each group separately. No significant differences were found between the two groups (p uncorrected > 0.05); sham and active groups were well matched by socio-demographic characteristics.

### Self-report data analysis

Self-report craving scores based on VAS (0-100) are visualized in *Figure 2*. Bar charts represent the mean value and error bars show standard error (SE) of the craving score at different assessment time points for each group separately. The LME showed a significant (p < 0.0001) effect of time, however, there was no statistically significant effect of group or time x group interaction. In pre-stimulation, we found that, in both groups, the craving score increased significantly (p uncorrected < 0.0001 in both groups) by fMRI cue induced craving task in the first scan (T2 (sham: 59.07 ± 6.17, active: 64.40 ± 5.20; Hedges’ g = 0.17, 95% confidence interval (CI) −0.35-0.69) compared to T1 (sham: 39.4 ± 6.07, active: 42.50 ± 5.26; Hedges’ g = 0.10, 95% CI −0.42-0.62); mean ± SE and effect sizes are reported). Our results showed that there is no significant increase (p uncorrected > 0.05) in craving self-report after the second exposure to the drug cues (T4 (sham: 24.67 ± 4.96, active: 27.23 ± 4.63; Hedges’ g = 0.17, 95% CI −0.35-0.69) compared to T3 (sham: 17.83 ± 21.55, active: 25.93 ± 3.77; Hedges’ g = 0.10, 95% CI −0.42-0.62); mean ± SE and effect sizes are reported). There was a significant (p uncorrected < 0.05 in both groups) reduction in craving after stimulation (T3 (sham: 17.83 ± 21.55, active: 25.93 ± 3.77) compared to T2 (sham: 59.07 ± 6.17, active: 64.40 ± 5.20); mean ± SE and effect sizes are reported) and between baseline (T0, the day before stimulation (sham: 49.37 ± 5.74, active: 61.37 ± 4.50; Hedges’ g = 0.42, 95% CI −0.10-0.94); and day after (sham: 11.17 ± 3.60, active: 12.67 ± 3.33; Hedges’ g = 0.08, 95% CI −0.44-0.60); mean ± SE and effect sizes are reported) stimulation in both groups. Results of DDQ data analysis and inside scanner craving can be found in *Figure S3* and *Figure S4* respectively. No significant effects of group or time by group interaction were found in DDQ.

**Figure 2:**
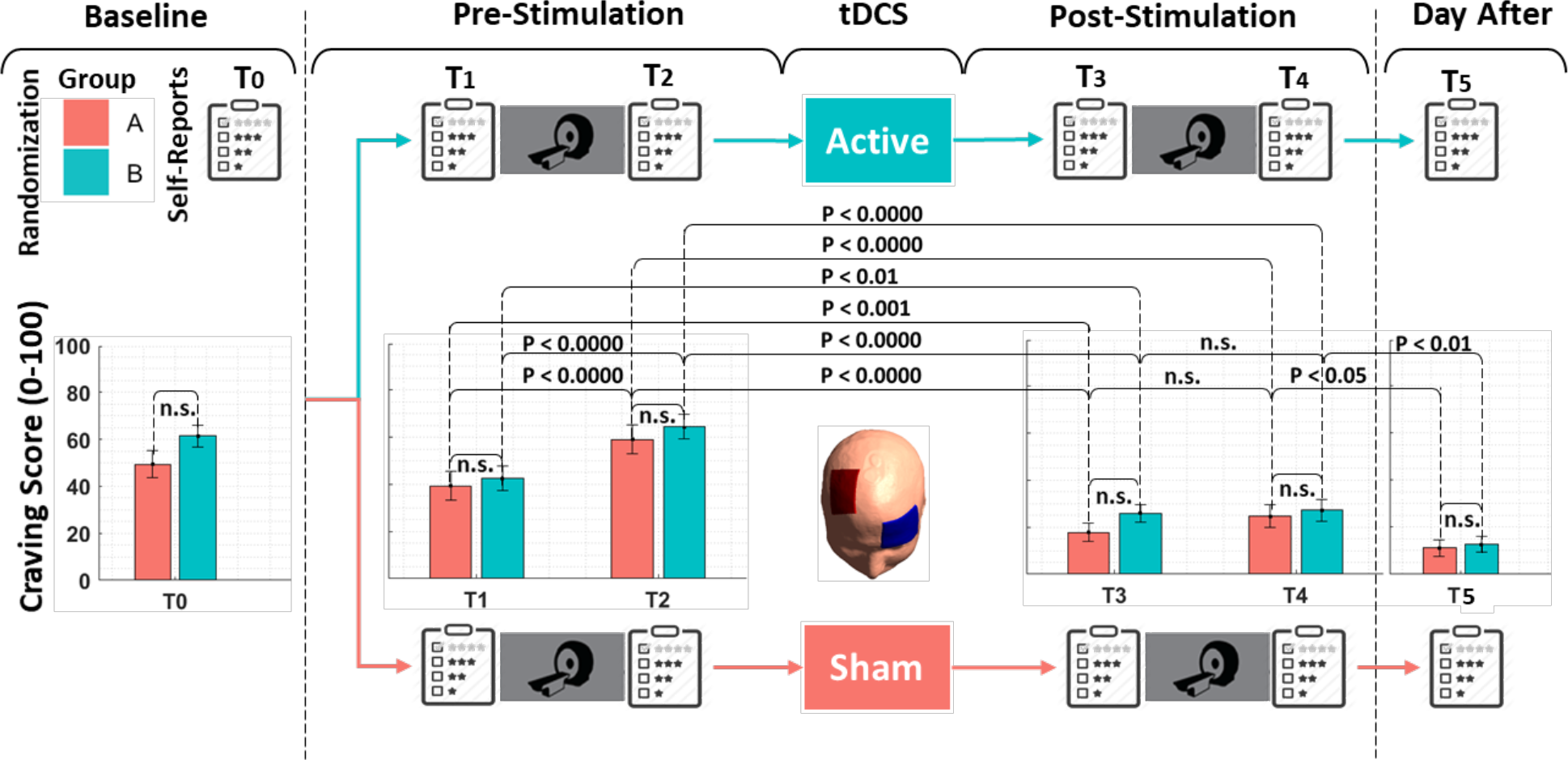
Craving assessment and results. Self-report craving scores based on VAS (0-100) were collected at 6 time points. Bar charts represent the mean value and error bars show SE of the craving score at assessment time for each group separately. Statistical results for between and within-group changes are shown above the bars based on uncorrected P values obtained from t-tests. As shown, there were no statistically significant differences in the craving score between active and sham groups. Except for T3 to T4, which showed no significant differences within each group, other time points demonstrated significant within-group differences. Abbreviation: VAS: Visual Analogue Scale, SE: standard error, n.s. non-significant.

In self-report and behavioral measures of response to the cues that are obtained inside the scanner, model free effect size showed Hedges’ g = −0.50 with 95% CI −1.03-0.03 (differences between meth and neutral blocks in post-stimulation compared to the pre-stimulation). LME model with group (active and sham) and time (first scan and second scan) as fixed effects, time x group as a mixed effect and subject as random effect showed a significant effect of time (p uncorrected < 0.0001). However, no significant effect of group or time x group interaction was found across the subjects. Post-hoc analysis showed that craving score was significantly different between meth and neutral cues in both groups in the first scan (p uncorrected < 0.0001; mean ± SE: sham group: meth = 2.907 ± 0.13, neutral = 1.692 ± 0.31; active group: meth = 3.209 ± 0.12,neutral = 1.707 ± 0.32), and was decreased significantly (p uncorrected < 0.01) in the second scan after both active and sham stimulation compared to the first scan (mean ± SE: sham group: meth = 2.353 ± 0.04, neutral = 1.379 ± 0.14; active group: meth = 2.306 ± 0.06, neutral = 1.415 ± 0.13).

For reaction time to craving, model free effect size for inside scanner craving (differences between meth and neutral blocks in post-stimulation compared to the pre-stimulation) showed Hedges’ g = 0.41 with 95% CI −0.11-0.94. LME model with time (first scan and second scan) and group (active and sham) as fixed effects, time x group interaction as mixed effect and subject as random effect, revealed significant effect of time (p uncorrected < 0.001) and no statistically significant effects of group or time by group interactions were found. Post-hoc analysis showed that reaction time (seconds) to the craving score, in the second scan (sham: meth = 1.600 ± 0.20, neutral = 1.442 ± 0.19; active: meth = 2.001 ± 0.29, neutral = 1.810 ± 0.27) compared to the first scan (sham: meth = 1.785 ± 0.19, neutral = 1.901 ± 0.31; active: meth = 1.839 ± 0.27, neutral = 2.223 ± 0.38), was significantly (p uncorrected < 0.0001) changed in response to the meth cues such that response time was increased in the active and decreased in the sham group.

We also checked the reaction time to a yellow box which was randomly appeared around one of the pictures in each block. Model free effect size for response to the yellow box showed Hedges’ g = 0.14 with 95% CI −0.42-0.7 (differences between meth and neutral blocks in post-stimulation compared to the pre-stimulation). LME model with time (first scan and second scan) and group (active and sham) as fixed effects, time x group interaction as mixed effect and subjects as random effects only showed a significant effect of time. Post-hoc analysis showed significant differences between meth (sham: 1.800 ± 0.42, active: 2.138 ± 0.18) and neutral (sham: 1.278 ± 0.20, active: 1.409 ± 0.14) blocks in both active (p uncorrected < 0.0001) and sham (p uncorrected < 0.01) groups during the first scan. In the second scan we did not find a significant effect of time (p uncorrected > 0.05). However, in the second cue exposure, a difference between meth (sham: 1.511 ± 0.27, active: 1.852 ± 0.26) and neutral (sham: 1.280 ± 0.39, active: 1.213 ± 0.21) blocks was significant (p uncorrected: sham > 0.05 and active < 0.0001) only in the active group with greater reaction time to the yellow box in the meth blocks. Behavioral and self-report measures of response to drug and neutral cues inside the scanner are depicted in *Figure S4*.

All of the 30 participants in each group tolerated the stimulation without any problem in their data collection. As reported in *Table 2*, there was no significant (p uncorrected > 0.05) difference between active and sham groups in terms of reported side effects. Severity of side effects and perceived relatedness to the stimulation can be found in supplementary materials (*Table S1*).

**Table 2:** Side effects (reported as 0 or 1) after post fMRI and day after stimulation session. Uncorrected p values are reported.

| Side effect | Total (n = 60) % (n) | Active % (n) | Sham % (n) | $P$ value <sup>a</sup> |
| --- | --- | --- | --- | --- |
| **After Post fMRI** |  |  |  |  |
| Headache | 5% (3) | 10% (3) | 0% (0) | 0.237 |
| Neck pain | 5% (3) | 10% (3) | 0% (0) | 0.237 |
| Scalp pain | 1.67% (1) | 3.33% (1) | 0% (0) | 1.000 |
| Tingling | 11.67% (7) | 13.33% (4) | 10% (3) | 1.000 |
| Itching | 5% (3) | 3.33% (1) | 6.67% (2) | 1.000 |
| Burning sensation | 8.33% (5) | 10% (3) | 6.67% (2) | 1.000 |
| Skin redness | 6.67% (4) | 6.67% (2) | 6.67% (2) | 1.000 |
| Sleepiness | 28.33% (17) | 33.33% (10) | 23.33% (7) | 0.568 |
| Trouble concentrating | 5% (3) | 6.67% (2) | 3.33% (1) | 1.000 |
| Acute mood change | 10% (6) | 13.33% (4) | 6.67% (2) | 0.671 |
| **Day After** |  |  |  |  |
| Headache | 6.67% (4) | 13.33% (4) | 0% (0) | 0.112 |
| Neck pain | 1.67% (1) | 3.33% (1) | 0% (0) | 1.000 |
| Scalp pain | 1.67% (1) | 0% (0) | 3.33% (1) | 1.000 |
| Tingling | 5% (3) | 3.33% (1) | 6.67% (2) | 1.000 |
| Itching | 3.33% (2) | 3.33% (1) | 3.33% (1) | 1.000 |
| Burning sensation | 0% (0) | 0% (0) | 0% (0) | 1.000 |
| Skin redness | 1.67% (1) | 0% (0) | 3.33% (1) | 1.000 |
| Sleepiness | 13.33% (8) | 16.67% (5) | 10% (3) | 0.707 |
| Trouble concentrating | 3.33% (2) | 3.33% (1) | 3.33% (1) | 1.000 |
| Acute mood change | 3.33% (2) | 0% (0) | 6.67% (2) | 0.492 |

<sup>a</sup> One-sided Fisher exact test

Results of the post-study questionnaire to determine the efficacy of blinding achieved by the strategy of this trial indicated that participants could not differentiate between the stimulation conditions (*Table 3*).

**Table 3:** Blinding efficacy assessed after the stimulation session and the day after.

| After Post fMRI |  |  |  |  |  |  | P value<sup>a,b</sup> |
| --- | --- | --- | --- | --- | --- | --- | --- |
| Was it 'Real' or 'Sham'? | Total (n = 60) |  | Active tDCS (n = 30) |  | Sham tDCS (n = 30) |  | 0.718 |
|  | Active%(n) | Sham%(n) | Active%(n) | Sham%(n) | Active%(n) | Sham%(n) |  |
|  | 85%(51) | 15%(9) | 83.3%(25) | 16.7%(5) | 86.7%(26) | 13.3%(4) |  |
| How much you are confident with your response? |  |  | Mean (SD)<br>78.87 (18.94) |  | Mean (SD)<br>78.03 (19.55) |  | 0.867 |
| Was stimulation effective to reduce craving? | Yes %(n) | No %(n) | Yes %(n) | No %(n) | Yes %(n) | No %(n) | 0.273 |
|  | 66.7%(40) | 33.3%(20) | 73.3%(22) | 26.7%(8) | 60%(18) | 40%(12) |  |
| How much this stimulation was effective (0-100)? |  |  | Mean (SD)<br>56.4 (28.68) |  | Mean (SD)<br>58.43(30.04) |  | 0.790 |
| Day After |  |  |  |  |  |  |  |
| Was it 'Real' or 'Sham'? | Active%(n) | Sham%(n) | Active%(n) | Sham%(n) | Active%(n) | Sham%(n) | 0.718 |
|  | 85%(51) | 15%(9) | 83.3%(25) | 16.7%(5) | 86.7%(26) | 13.3%(4) |  |
| How much you are confident with your response? |  |  | Mean (SD)<br>75.97 (21.60) |  | Mean (SD)<br>76.2 (24.67) |  | 0.969 |
| Was stimulation effective to reduce craving? | Yes %(n) | No %(n) | Yes %(n) | No %(n) | Yes %(n) | No %(n) | 0.559 |
|  | 73.3%(44) | 26.7(16) | 76.7%(23) | 23.3%(7) | 70%(21) | 30%(9) |  |
| How much this stimulation was effective (0-100)? |  |  | Mean (SD)<br>56.50(31.04) |  | Mean (SD)<br>61.23(25.67) |  | 0.522 |

<sup>a</sup> Chi-Square Tests, <sup>b</sup> Independent-Samples t-test

### Task-based functional activity results

Active clusters obtained from whole-brain functional activity analysis with significant time (post vs pre) by group (active vs sham) interaction are visualized over 3D standard MNI space. As illustrated in *Figure 3*, five active clusters in the left hemisphere survived our predefined threshold (voxel-level threshold: p uncorrected < 0.005, cluster size > 40); medial frontal gyrus (MFG), anterior insula (Ins), inferior parietal lobule (IPL), precuneus, inferior frontal gyrus (IFG). The precise location of each cluster as well as number of active voxels are reported in *Table 4*.

**Figure 3:**
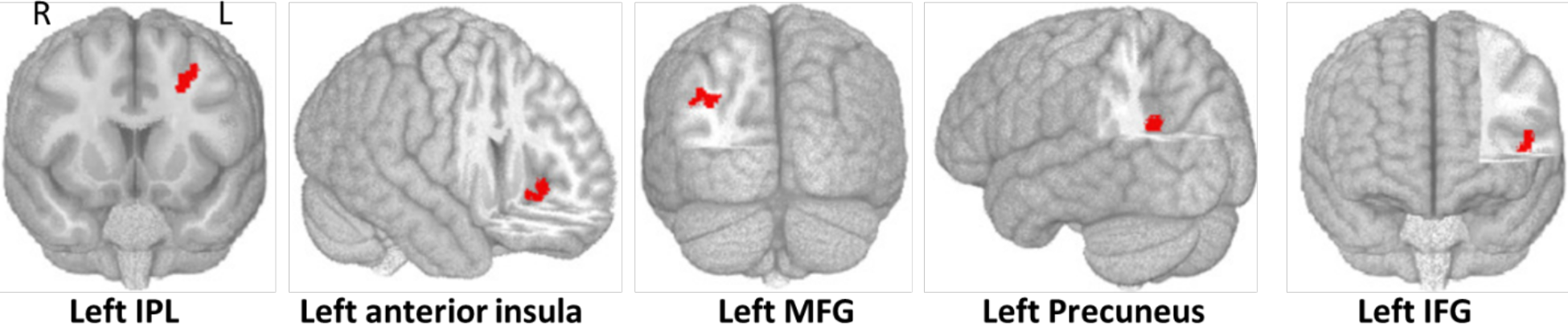
Time by group interaction during fMRI cue-reactivity task. Active clusters (voxel-level threshold: p uncorrected < 0.05 and cluster level threshold: cluster size < 40) obtained from LME models (with time x group interaction as the fixed effect and subject as random effect; time: post vs pre-stimulation, and group: active vs sham) that showed significant time by group interaction. From left to right cluster location in MNI space: MFG (−53, −87, 33) with 105 voxels, anterior insula (−27 19 39) with 88 voxels, IPL (−35 21 −7) with 87 voxels, precuneus (−53 15 9) with 49 voxels, and IFG (−19, −63, 25) with 47 voxels. Abbreviation: MFG: medial frontal gyrus, IPL: inferior parietal lobule, IFG: inferior frontal gyrus.

**Table 4:** Functional activity analysis results. Task-modulated clusters during the fMRI cue-reactivity task with significant time by group interaction. Voxel-level threshold: p uncorrected < 0.005 and cluster-level threshold: cluster size < 40. Abbreviation: MFG: medial frontal gyrus, IPL: inferior parietal lobule, IFG: inferior frontal gyrus.

| Cluster location | Cluster size (# voxels) | Cluster volume (mm<sup>3</sup>) | Peak coordinate |  |  | F-value in peak |
| --- | --- | --- | --- | --- | --- | --- |
|  |  |  | X | Y | Z |  |
| Left IPL | 105 | 1050 | -53 | -87 | 33 | 17.58 |
| Left Anterior Ins | 88 | 880 | -27 | 19 | 39 | 21.31 |
| Left MFG | 87 | 870 | -35 | 21 | -7 | 18.61 |
| Left Precuneus | 49 | 490 | -53 | 15 | 9 | 15.92 |
| Left IFG | 47 | 470 | -19 | -63 | 25 | 21.88 |

Averaged EFs within significant clusters were extracted from individualized CHMs transformed to the MNI space. Using regression models (alteration in functional activity (post minus pre) within each cluster as the dependent variable, and group (as well as induced EFs within the cluster as independent variables), we found no significant effect for EFs but the effect of group was significant (p uncorrected < 0.000) for all above-mentioned clusters. Post-hoc analysis of cluster-based functional activity in pre- and post-stimulation and scatter plots for determining the correlation between functional changes and induced EFs at the cluster-level are visualized in *Figure S5* for each group separately (although the injection of current had stopped after 40 sec of active stimulation in sham stimulation, EF values in this group are hypothetical based on the computational head model for active stimulation). We also checked for the correlation between cluster-based functional activity and changes in the craving in both groups. However, no significant correlation was found.

Based on our exploratory approach, functional activity analysis results using BNA parcellation are visualized in *Figure 4* for sham and active groups—signal change (post minus pre). The level of brain activation during the task was extracted for each group separately at each time point (pre- and post-stimulation). In *Figure 4*, changes in brain activation are depicted for sham (in red; panel a) and active (in blue, panel b). There is a wide-spread habituation in response to drug cues in the second exposure (second fMRI scan after intervention) in the sham group (negative direction in panel a). In contrast, as shown in panel b, there are areas with significant increase in their activation in response to the active stimulation. The coefficient of time by group interaction term in LME models are also reported for all BNA regions across the population in panel c and brain regions with significant (p uncorrected < 0.05) time x group interactions are also determined with the color purple. None of the regions survived FDR correction.

**Figure 4.**
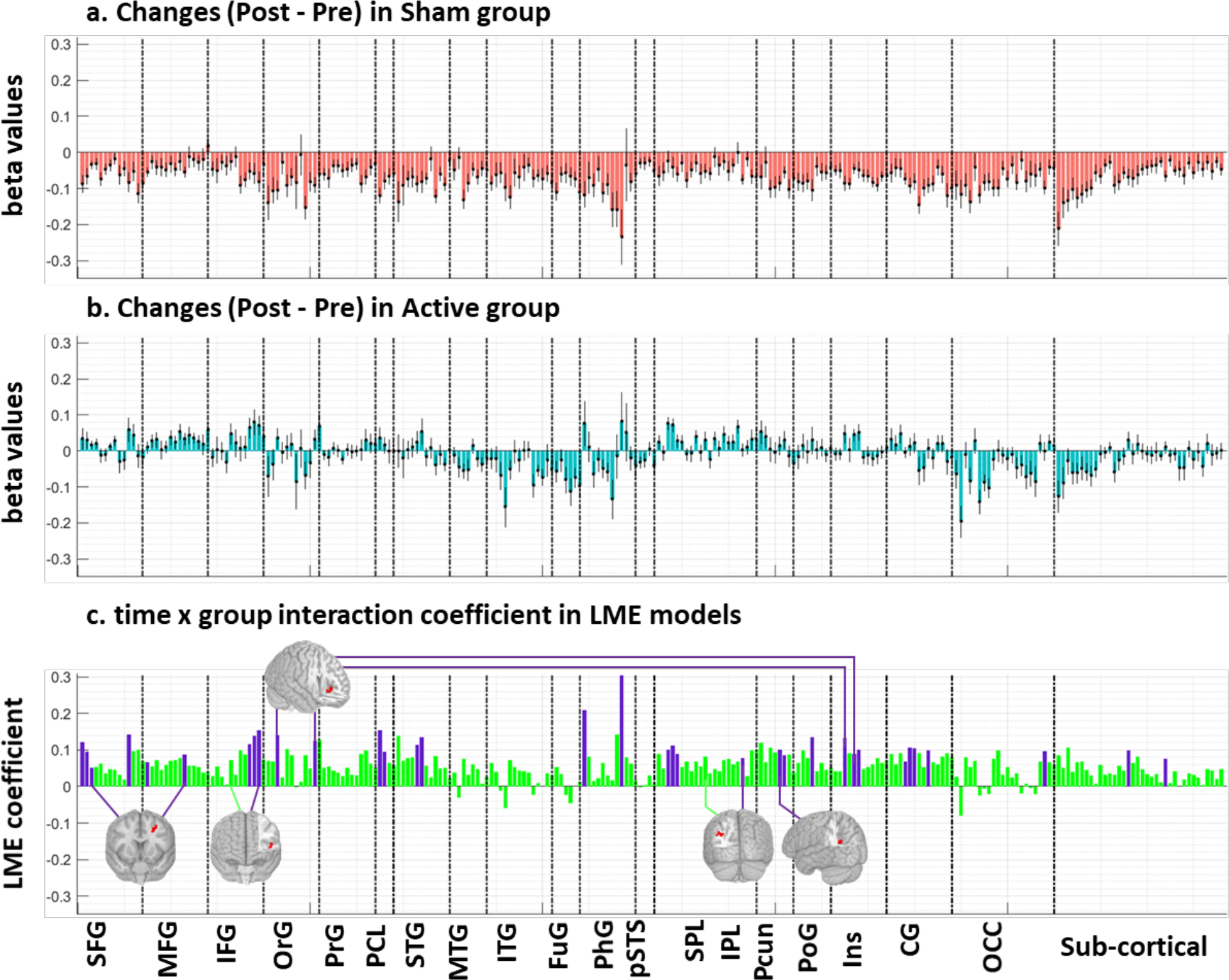
Change in brain activation during task-based fMRI in Brainnetome atlas (BNA) parcellation. In an exploratory approach, brain activation during the methamphetamine cue reactivity (MCR) task was extracted in pre- and post-stimulation fMRI data collection. Based on whole-brain analysis, changes in brain activation (Post minus Pre) in terms of beta values obtained from the general linear modeling (GLM) are represented for the sham group (**Panel a**) and active group (**Panel b**) to show the direction of changes in functional activity in each group separately. Bars show mean value and error bars show standard error of the beta values across the population in each group. As shown in the first row, a negative direction (Post < Pre) was found in all BNA subregions in sham stimulation. In active stimulation, both positive and negative directions were detected. **Panel c.** Time x group interaction coefficient in linear mixed effect (LME) models: Bars show the coefficient of the time by group interaction term in an LME (time x group as fixed effect and subjects as a random effect) model for each subregion in BNA. Brain regions with significant time by group interaction term (P uncorrected < 0.05) are shown in purple. 3D brains are active clusters in whole-brain analysis depicted in **Figure 3**. BNA subregions that overlap with active clusters were determined by connecting a line between BNA areas and related active clusters. Abbreviation: LME: linear mixed effect model, SFG: superior frontal gyrus, MFG: middle frontal gyrus, IFG: inferior frontal gyrus, OrG: orbital gyrus, PrG: precentral gyrus, PCL: paracentral lobule, STG: superior temporal gyrus, MTG: middle temporal gyrus, ITG: inferior temporal gyrus, FuG: fusiform gyrus, PhG: parahippocampal gyrus, pSTS: posterior superior temporal sulcus, SPL: superior parietal lobule, IPL: inferior parietal lobule, Pcun: precuneus, PoG: postcentral gyrus, Ins: insula, CG: cingulate, OCC: occipital cortex.

### Task-modulated functional connectivity results

CHMs generated for all participants and personalized head models in native space can be found in *Figure S6*. Head models were transformed to the standard fsaverage space for group-level analysis. The mean and standard error of the EFs were extracted from all BNA subregions. As schematically shown in *Figure 5*, parcellation of CHMs using BNA showed that lateral area of the SFG in BNA sub-regions (SFG6; mean ± SD = 0.2772 ± 0.05) received the highest averaged EFs across the population.

**Figure 5:**
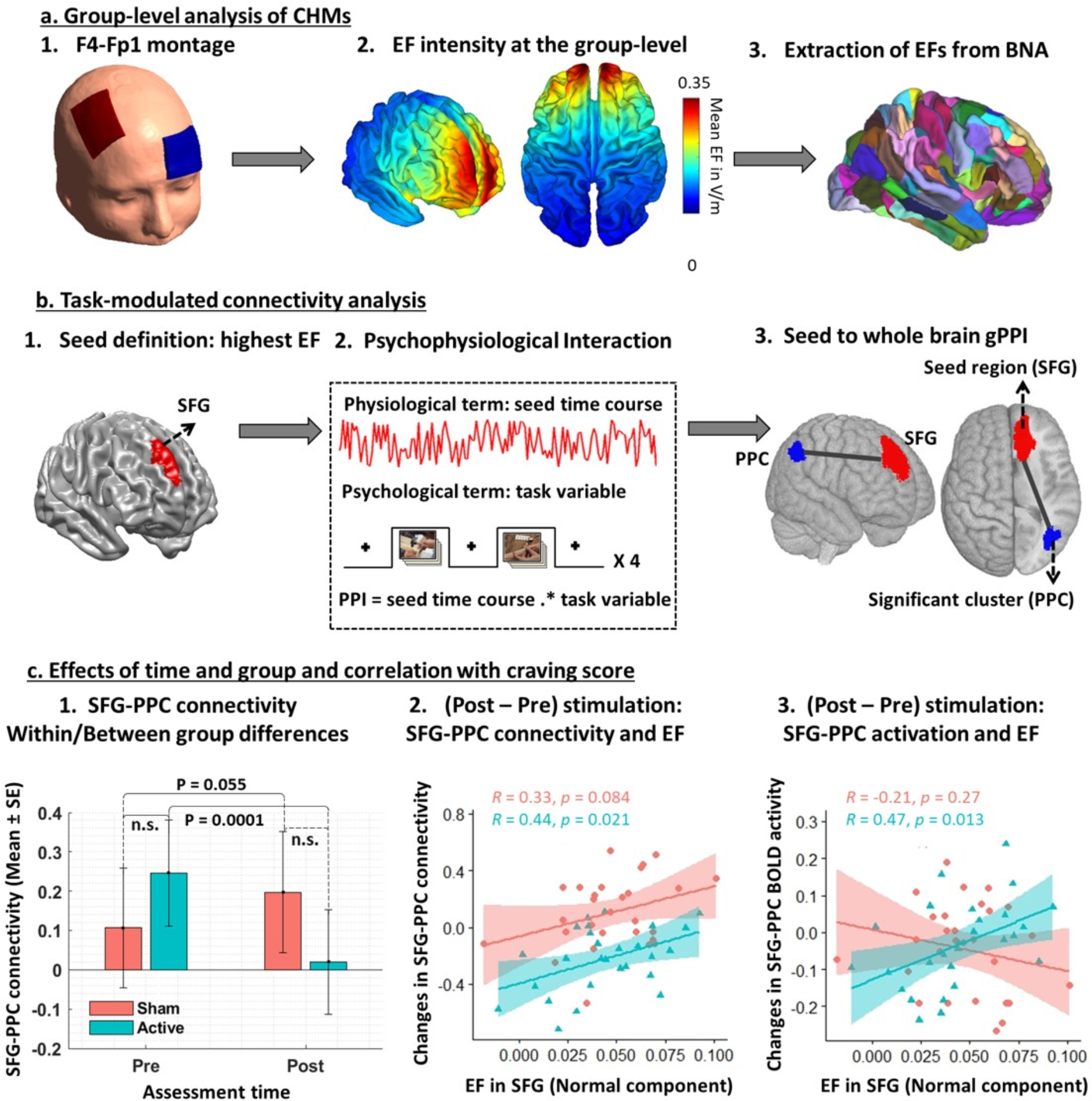
Task-modulated (cue reactivity) connectivity analysis: computational head model (CHM) approach. a. Group-level analysis of CHMs: 1. Anode/cathode electrodes were placed over F4/Fp1 location in EEG standard system, 2. Individualized computational head models (CHMs) were generated for all 60 participants to calculate EF distribution patterns using finite element modeling. Then, CHMs were transformed to fsaverage standard space for group-level analysis, 3. Brainnetome atlas (BNA) was applied to head models in standard space and averaged EFs were calculated for each region. **b. Task-modulated connectivity analysis:** 1. The BNA region with the highest averaged EF (lateral part of right SFG) was selected as a seed region for gPPI analysis. 2. BOLD signal was extracted from the seed region (in red) and clusters with significant (voxel-level threshold at p uncorrected < 0.001 and the cluster-level threshold at p FDR corrected < 0.05) time x group interaction is indicated over a 3D brain (in blue). 3. Mean values with error bars representing the connectivity between SFG and task-modulated cluster (which is named PPC here) in pre- and post-stimulation fMRI scans. **c. Effect of time and group and correlation with EF.** First column: Bars show mean and error bars show standard error in SFG-PPC connectivity for each group at each time point with a significant effect of time in the active group. Post-hoc results are reported above bar charts. Second column: Scatter plot for the correlation between the normal component of the EF in SFG and changes in SFG-PPC task-modulated connectivity. Third column: Scatter plot for correlation between the normal component of the EF in SFG and changes in SFG BOLD signal activation. Pearson correlation coefficients and p values for each group are reported above scatterplots. Abbreviation: EF: electric field, BNA: Brainnetome atlas, gPPI: generalized psychophysiological interaction, SFG: superior frontal gyrus, n.s.: non-significant, SE: standard error.

The SFG area that received the highest electrical current was used as the seed for whole-brain task-modulated connectivity analysis (gPPI). By considering p uncorrected < 0.001 at the voxel level and p FDR corrected < 0.05 at the cluster level, we found one significant cluster in the right hemisphere in which task-modulated connectivity with the frontal seed showed a significant time (post vs pre) by group (active vs sham) interaction. This cluster, which is shown in *Figure 5.b*, is centered in (36, −60, 38) as (x, y, z) in MNI space and contains 330 voxels. Our results demonstrated that task modulated connectivity between the frontal seed and a significant cluster in the parietal cortex (posterior parietal cortex (PPC) in *Figure 5.b*) decreased in the active group in the second scan (first scan: 0.25 ± 0.14, second scan: 0.02 ± 0.13, mean ± SE). On the contrary, SFG-PPC increased in the sham group in the second scan compared to the first one (first scan: 0.11 ± 0.15, second scan: 0.20 ± 0.15; mean ± SE).

As shown in *Figure 5.c*, the normal component of the EF in the SFG was extracted from CHMs. In an exploratory analysis, the correlation between normal EF in SFG and changes (post - pre) in SFG-PPC connectivity was calculated for each group separately and a significant correlation was found only in the active group (R = 0.44, p = 0.021). Changes in (post – pre) BOLD signal functional activity in the SFG was also calculated and showed significant correlation with the normal component of the EF in the SFG only in the active group (R = 0.47, p = 0.013). However, a significant correlation between tangential EF in the SFG and functional activity connectivity related to the SFG was not found.

## Discussion

This pre-registered, triple-blind, randomized, sham-controlled clinical trial examining the effects of tDCS in response to drug cue exposure in a group of individuals with methamphetamine use disorder yielded six main results. First, based on self-report, both active and sham groups experienced a significant reduction in craving when comparing pre- vs. post-stimulation and the day after stimulation. However, there was no significant main effect of active stimulation vs sham stimulation in reducing craving. Second, there was widespread habituation (decreased functional activity) to drug-related cues in the sham group during the post-stimulation scan. In comparison, a number of brain areas showed increase in functional activity during the second cue reactivity task in the active stimulation group. Third, whole brain fMRI analysis with LME revealed five main clusters with significant positive (higher activation in the active stimulation group) time by group interaction; medial frontal gyrus, anterior insula, inferior parietal lobule, precuneus, and inferior frontal gyrus—all clusters located in the left hemisphere. Fourth, we simulated individualized computational head models (CHMs) and found a significant effect of group in the relationship between level of current in the above-mentioned significant clusters and changes in task-modulated activation. Fifth, based on a seed-to-whole brain psychophysiological interaction analysis and CHMs at the individual level, we found that the brain region with the highest averaged electric field (right superior frontal gyrus) had significant time by group interaction with the posterior parietal cortex. Finally, this frontoparietal task-modulated connectivity, which was decreased in the active group after application of tDCS, showed a significant positive correlation with the normal component of the EFs within brain regions that had the highest EFs.

### Stimulation effects on subjective response: craving score

Irrespective of active or sham stimulation, subjective response to drug cue exposure showed significant increases in craving after the first scan (cue exposure) (pre-stimulation, T2 compared to T1). The craving score decreased significantly after the tDCS intervention in both groups (T3 compared to T2). However, there was no significant change in craving after the second scan (cue exposure) (post-stimulation, T4 compared to T3) in either the active or sham groups (lack of subjective response to drug cue in the second exposure). There was a significant reduction in the craving self-report from baseline (before the stimulation day) to the day after stimulation without any significant difference between groups. Our findings suggest that our interventions, which included both cue exposure and active/sham tDCS, successfully reduced craving in both groups. However, no significant between-group differences or time x group interaction was found across the population. There have been other tDCS studies that reported no significant differences between active and sham groups in terms of self-reported drug craving (see this systematic review (Lupi et al. 2017)). For instance, Xu et al. (Xu et al. 2013) reported that reduction in cigarette craving after bilateral DLPFC stimulation did not differ significantly between active and sham stimulation. In another randomized sham-controlled trial, craving scores did not change significantly by applying a multi-session bilateral DLPFC stimulation in a group of people with severe alcoholism (Klauss et al. 2014). Furthermore, five sessions of unilateral tDCS over DLPFC (F3-Fp2) in a group of participants with schizophrenia showed no statistically significant effects on cigarette craving or cigarettes smoked (Smith et al. 2015). Another study among cigarette smokers on the immediate effects of unilateral DLPFC tDCS during an *in vivo* smoking cue exposure, revealed no significant time (pre vs post) by group (sham vs active) interaction on self-reported craving (Kroczek et al. 2016). Meanwhile, there have been several studies among people with SUDs that reported significant differences between and sham groups in terms of drug craving (cocaine (Batista et al. 2015), marijuana (Boggio et al. 2010), heroin (Sharifi-Fardshad et al. 2018), alcohol (Klauss et al. 2018), methamphetamine (Shahbabaie et al. 2018)). Heterogeneity in methodological details and large inter-individual variabilities in response to tDCS can contribute significantly to the mixed results (Bashir and Yoo 2016). The inconsistency between different studies might be attributed to the differences in the number of subjects, type of substances, duration of abstinence, and state of dependency in the target population and methodological details like electrode montage and sham stimulation protocols and details in cue exposure protocols. For example, compared to the previous tDCS-fMRI research in the field of addiction medicine, we used a larger sample size in this study. Significant between-group differences in behavioral outcomes reported in previous studies might be related to the smaller sample size and low statistical power that reduces the likelihood that statistical results reflect a true effect (Button et al. 2013) as studies with smaller size are more likely to report false positives with relatively large effect size (Minarik et al. 2016).

### Stimulation effects on neural response: functional activity

Cue-induced craving, which is used in this study, is one of the most frequently used ecologically valid paradigms to induce craving and measure the effects of interventions on drug cue exposure (Ekhtiari, Zare-Bidoky, et al. 2020). With respect to the neural substrates of cue-reactivity in participants with SUDs (Hanlon et al. 2018), we identified several key brain regions related to cue-reactivity including medial frontal gyrus, anterior insula, inferior parietal lobule, precuneus, and inferior frontal gyrus that showed significant time by group interactions. Our results suggest that active tDCS compared to sham modulates brain activity both locally (near the stimulating site; e.g., MFG) and in regions distant from the stimulating electrodes such as IPL or precuneus. The overall pattern of activation in post-hoc analysis showed that, in contrast to sham stimulation, active tDCS increases functional activity in the above-mentioned clusters. This may reflect differential modulatory input or output from these areas during cue exposure after active stimulation. This modulatory effect can be interpreted by the induced electrical field (EFs) at the individual level within these clusters in the active group as our EF simulation results confirm significant effect of group in the relationship between change in functional activity and tDCS-induced EFs.

Even if applied locally, considering large-scale network connectivity and with respect to the diffusivity of the current in conventional tDCS, tDCS likely influences the neuronal activation in various parts of the brain directly or indirectly (Ghobadi-Azbari et al. 2020). Our exploratory approach showed that during the second cue exposure (second fMRI scan after intervention) there are areas with significant increased activation after active stimulation. In contrast, a widespread decrease in activations in response to the second exposure to drug cues (second scan) was found in the sham group. Irrespective of the mechanism, the observation of widespread decreased activation in the second exposure to drug cues after the sham stimulation while active tDCS was associated with increased functional activity in some specific brain regions, lead us to hypothesize that tDCS may effectively modulate brain response to drug cues. However, the subjective self-reports of craving do not support this modulation having an immediate effect on self-reported craving.

### Stimulation effects on neural response: task-modulated connectivity

In addition to task-based functional activity, recently, the use of task-based fMRI has become critical in probing tDCS induced changes in connectivity modulation, which may be especially advantageous in a context-dependent analysis (Sehatpour et al. 2020, Li et al. 2019, Ganho-Ávila et al. 2019, Wu et al. 2015). Task-modulated connectivity suggests that functional connectivity may largely depend on the task that is performed during the application of tDCS. Here, we found tDCS induced a significant reduction of frontoparietal task-modulated connectivity in the active stimulation group while this connectivity was increased among participants in the sham group with a trend towards significance (p uncorrected = 0.055) in post-hoc analyses. Our results suggest that conventional tDCS targeting DLPFC may act upon decoupling of the frontal target and its ipsilateral connectivity with the parietal cortex in the context of cue exposure. Task-modulated connectivity alterations due to effects of stimulation may have several reasons, such as the existence of excitatory/inhibitory functional connections between two regions or the presence of intermediate nodes that are dependent on the seed region. The seed region in this study was located in the right superior frontal gyrus where our simulations predicted the strongest averaged EF intensity for the F4-Fp1 electrode montage across the population in this brain region. Significant correlations were found between EFs (normal component) within SFG and functional activity/connectivity related to this brain region. This demonstrated the usefulness of CHMs to inform regional data analysis in neuroimaging studies. Furthermore, it also suggested that stimulation effects could be extended to distant brain regions where there is stronger impact on brain regions that are spatially distributed but functionally connected.

### Absence of between group differences in subjective response despite significant neural changes

Finally, it is important to highlight that we found significant tDCS neural activity as well as connectivity effects in the absence of between-group differences in self-reported craving. The absence of between-group differences in self-reported craving leads to hypotheses about potential causes. There are several reasons why group differences in self-reported data between the active and sham stimulation groups may not have been found in our study when they have been observed in many previous tDCS studies in the field of SUDs. Four potential reasons are listed in the following paragraphs.
1. **Sham protocol:** With our version of the NeuroConn DC-stimulator MR, sham stimulation consisted of delivering an active stimulation for 40 sec to mimic the sensations observed with active tDCS. Electrode-to-skin impedance was checked every 15 sec by injecting low current intensity. Previous studies investigated tDCS effects with parameters similar to the sham stimulation with short duration of active stimulation and reported that sham protocols with short active stimulation may induce direct neurobiological effects measurably different from 0 mA stimulation (Javadi, Cheng, and Walsh 2012, Furubayashi et al. 2008, Fonteneau et al. 2019, Nikolin et al. 2018).
2. **Placebo effects:** Our results indicated that more than 70% of participants in each group thought that the stimulation was effective in reducing craving. Therefore, part of stimulation outcomes may be induced by a strong placebo effect (Fonteneau et al. 2019). tDCS, as a physically sensible intervention, can trigger expectations (positive or negative) and may affect overall outcomes (Supino 2012). Therefore, it is important to subtract the placebo effects caused by sham stimulation from active effects induced by active stimulation to determine the true effect induced by tDCS alone. However, the parallel design used in this study with only two arms for active and sham stimulation does not allow for discriminating such placebo effects (Wörsching et al. 2017).
3. **Complex interactions between stimulated brain regions:** There might be a complex interaction between different neural responses to the stimulation and final self-report outcomes within our cue exposure protocol. The cue reactivity task in the first scan was effective in increasing functional activity in certain brain regions in response to drug cues. Previous findings support tDCS interactions with concurrent brain activity, preferentially modulating brain regions or networks that are already activated (Boroda et al. 2020, Bikson and Rahman 2013). Hence, with respect to the activity-specificity and diffusivity of the current in conventional tDCS, our intervention modulated different parts of the brain with cumulative impact of neural stimulation through inhibitory/excitatory pathways that eventually did not reveal significant change in subjective outcomes.
4. **Role of cue exposure:** Participants were exposed to 24 meth cues in the first fMRI scan before stimulation. The response to drug cues was reflected in both an increase in craving self-report outside the scanner before imaging to after imaging and also behavioral and subjective self-reports inside the scanner (Figure 2, S3 and S4). However, in the second scan (cue exposure) these effects were reduced or non-existent in both groups who were actively involved in their recovery process in the first few weeks of their abstinence. There is a risk that a very powerful habituation response to drug cues created a ceiling effect on the actual effect of tDCS which is probably smaller and more variable among participants.

### Methodological strengths and recommendations for future tDCS clinical trials

In this study, specific methods were employed that could increase transparency and replicability in future tDCS studies SUDs. **1) Triple blinding:** In our study, research participants, research staff who administered the stimulation and performed assessments, and individuals who did the data analysis and assessed the outcomes did not know which intervention (active or sham) was administered. Despite the greater complexity, triple-blinding is recommended for tDCS studies in order to reduce assessment or analysis bias and increase accuracy and objectivity of stimulation outcomes. **2) Accurately reporting sham procedures:** In the methods section, we prepared a detailed definition of our sham procedure and timing charts of sham stimulation. Previous findings confirmed that inconsistent results across different sham-controlled clinical trials might arise from sham inconsistencies (Fonteneau et al. 2019, Neri et al. 2020). In tDCS research, conclusions are drawn based on comparisons between active and sham groups making it critical to report the type of stimulation device and sham timing chart used. This practice will increase the reproducibility of tDCS studies and make results comparable across different studies. **3) Reporting time of stimulation during the day:** In our study, all participants received stimulation in the afternoon and we accurately reported the time of start of stimulation for each individual to make sure both groups received stimulation in the same diurnal context (Figure S1). As suggested in previous studies, it is crucial to pay attention to stimulating research participants at a similar time of day. Reporting this factor can facilitate outcomes that are replicable and may help to reduce inter-individual variability (Salehinejad, Kuo, and Nitsche 2019, Sale, Ridding, and Nordstrom 2007). **4) Reporting overall brain response:** Atlas-based parcellation of functional activity to look at overall changes through the brain in each group separately helped us to find an interesting trend in the direction of changes after sham stimulation (Figure 4) that could not be easily inferred by typical whole-brain thresholding analysis. This type of visualization might be informative for future tDCS studies to determine overall effects of stimulation. **5) Integrating electric field modeling with neuroimaging data:** Using CHMs to inform fMRI analysis helped us to determine the relationship between highly modulated brain regions and other parts of the brain based on task modulated activity and connectivity. Furthermore, integrating accurate electric field models with neuroimaging data can help to capture inter-individual variability of the EFs across participants. tDCS-induced EFs based on personalized CHMs might be informative in investigating the impact of EF differences on tDCS neural and behavioral outcomes. This may help strengthen the understanding of the contribution between tDCS-induced EFs and the neural mechanism of action for each individual and across the study population.

### Limitations and future directions

As with all neuroimaging/neuromodulation studies, there are many potential limitations and confounds that need to be considered in interpreting this study. There is not enough experimental evidence to make solid speculation regarding the relationship between the effects of induced EFs, the appearance of BOLD signals, and changes in self-reported craving. In this study, we only checked for a linear relationship between these factors (based on using linear mixed effect models, linear regression, and correlation). These three factors might not have a simple linear relationship, however. More complex models might be needed to measure relationships between different stimulation outcomes.

A reason for the lack of significant differences in subjective reports in this study may be related to our parallel experiment design that compares two distinct groups of participants with each other with a wide range of interindividual variation. Previous findings suggest that paired analysis in crossover designs provides better power than an unpaired one and therefore, in future studies, crossover studies might be a better design for comparing equivalent intervention than parallel-group studies because participants serve as their own control (Cleophas and de Vogel 1998). Furthermore, under the assumption that active and placebo effects are additive, despite a parallel design, crossover studies would make it possible to subtract placebo and nocebo effects from overall responses to obtain active stimulation effects for each individual (Enck, Klosterhalfen, and Zipfel 2011).

In this study, we only focused on unilateral tDCS over the right DLPFC with two large electrode pads. Inspired by a previous TMS-fMRI study in the field of depression (Cash et al. 2020), rather than anatomical targeting and “one-size-fits-all,” stimulation dose, it might be important to propose a customized MRI guided multi-electrode montage for each participant based on individualized brain activity/connectivity at baseline and target specific brain functions related to cue-induced craving as suggested for each person (Hanlon et al. 2018).

In our pre/post tDCS-fMRI design, neural response showed habituation to post-stimulation cue exposure that can affect other stimulation outcomes such as subjective reports. In order to reduce habituation effects during cue exposure, instead of offline fMRI data collection immediately before and after stimulation, it might be better to use post-stimulation only fMRI or concurrent tDCS-fMRI during a cue exposure task to increase the efficacy of task-modulated outcomes. Furthermore, online tDCS-fMRI trials can help to close the loop between ongoing brain-state and stimulation parameters (e.g., current intensity, or electrode montage, optimal number and duration of the sessions) for each individual to optimize stimulation outcomes by tailoring the intervention to better fit each individual (Ghobadi-Azbari et al. 2020).

## Conclusion

Our study confirms that tDCS is a safe, non-invasive technique with minimal adverse effects in a group of participants with MUDs. The blinding was effective while there was a strong effect for sham stimulation on the perceived effectiveness of stimulation on drug craving without significant difference with active stimulation. Non-significant between-group differences in craving scores with decreased craving in both groups suggested a complex interaction between tDCS effects and habituation of cue effects that may eventually affect self-reported outcome measures. Significant time by group interactions in task-based fMRI data with increased functional activity only in the active stimulation group with a significant role of tDCS-induced EFs at the individual level suggests a modulatory role of tDCS during cue exposure. However, more research is needed to determine the relationship between neural activation and craving induced by drug related cues. We have also showed that task-based connectivity between highly modulated brain regions and other parts of the brain can be changed by tDCS. This alteration in task-modulated connectivity has a significant correlation with craving score. These results suggest that integrating tDCS with fMRI drug cue reactivity can be used in future studies to understand how tDCS affects neural processes underlying drug craving in people with MUDs and how correlation between induced EFs and neural response (activity as well as connectivity) can be used as a predictor of behavioral outcomes such as craving score. However, there are still many questions on how the neuromodulatory effect of tDCS can be consistently translated to a clinical application.

## Data Availability

The data and analysis codes that support the findings of this study are available from the corresponding author upon reasonable request. The fMRI task and its codes are available in https://github.com/rkuplicki/LIBR_FDCR_Dynamic.

## Data availability

The raw database and CHMs generated for this study are available on request to the corresponding author.

---

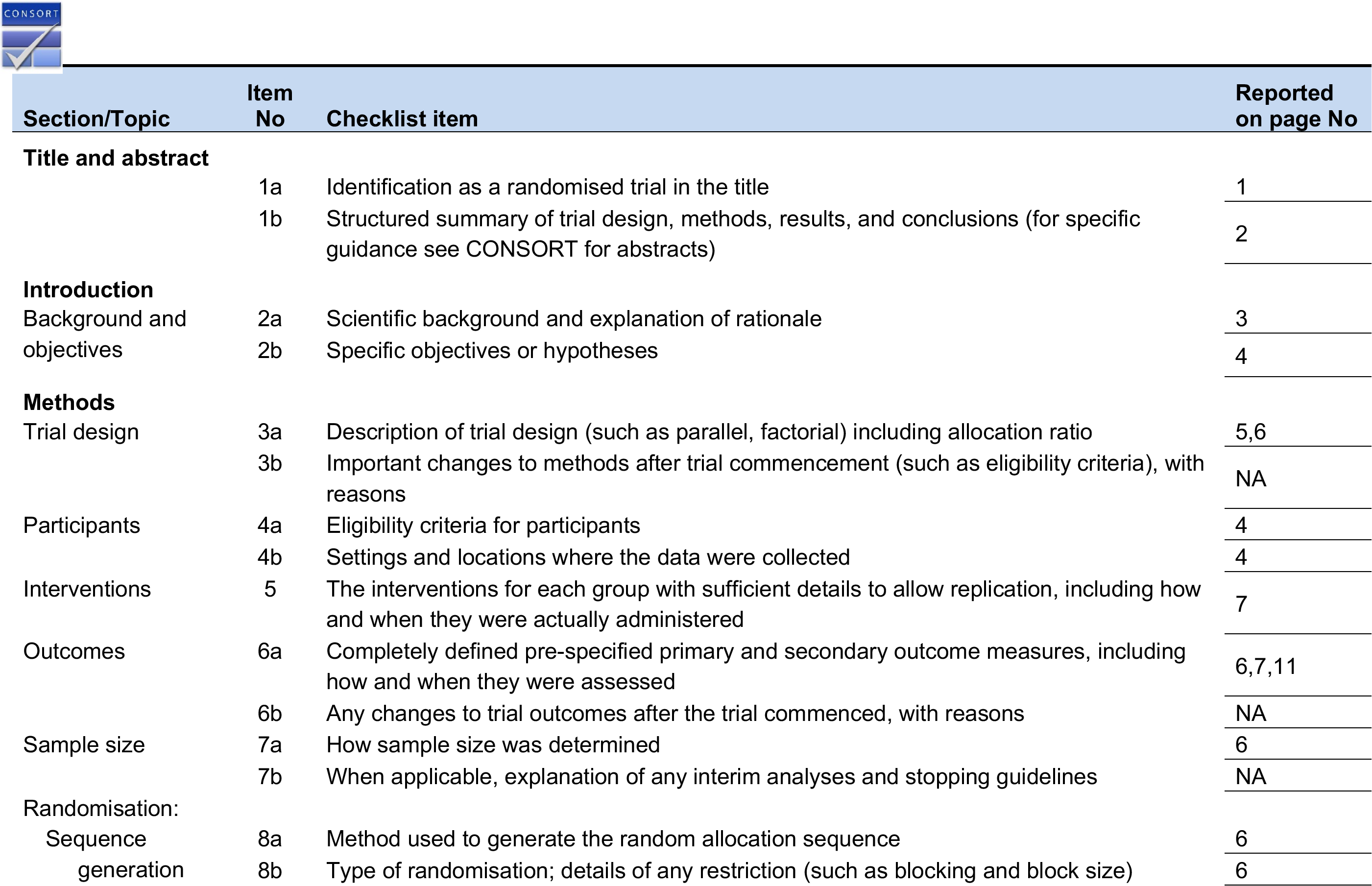

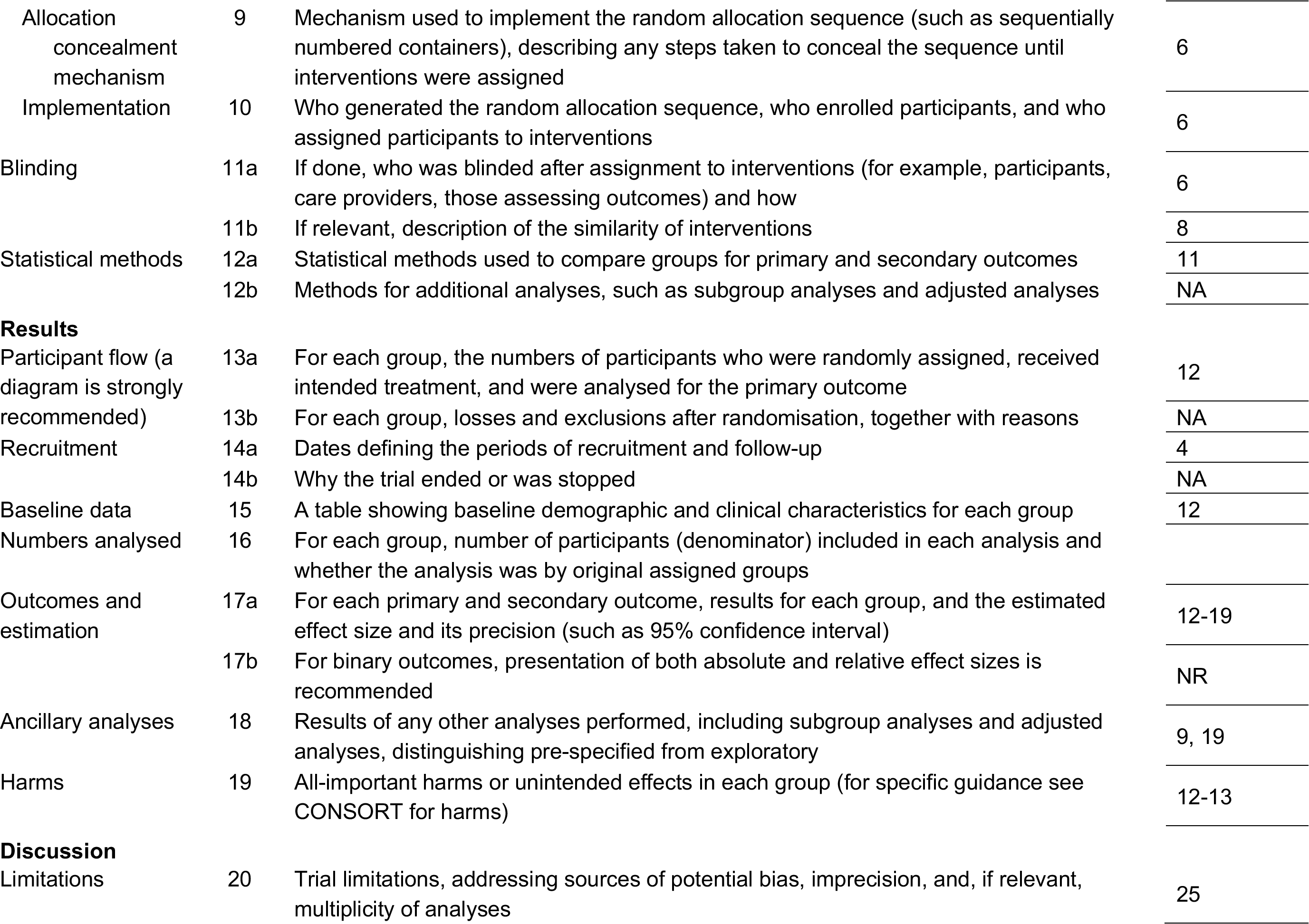

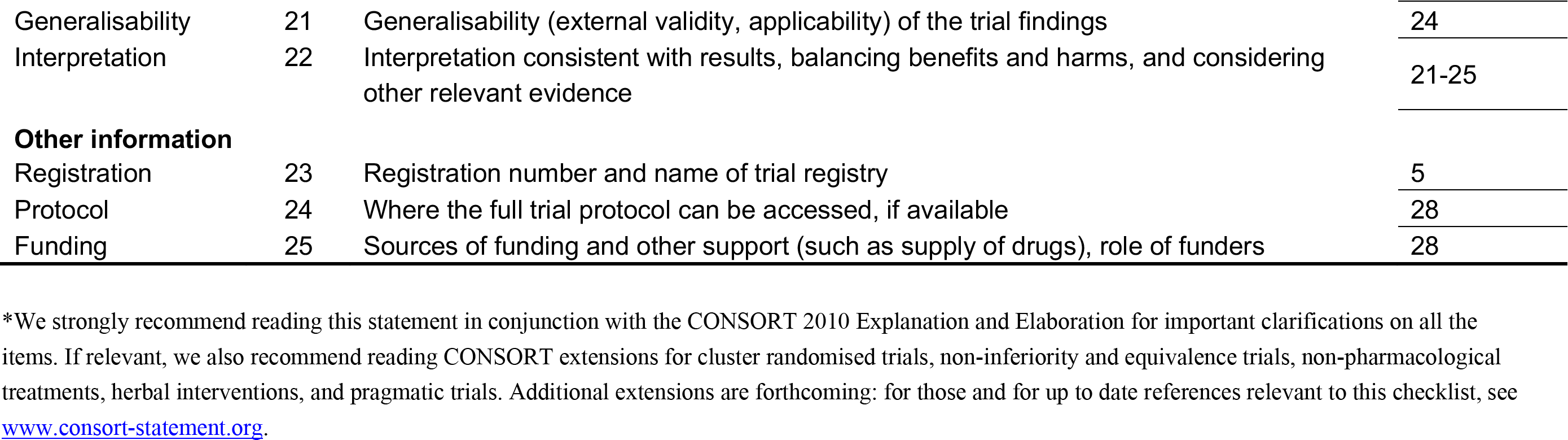
CONSORT 2010 checklist of information to include when reporting a randomised tria^*^.

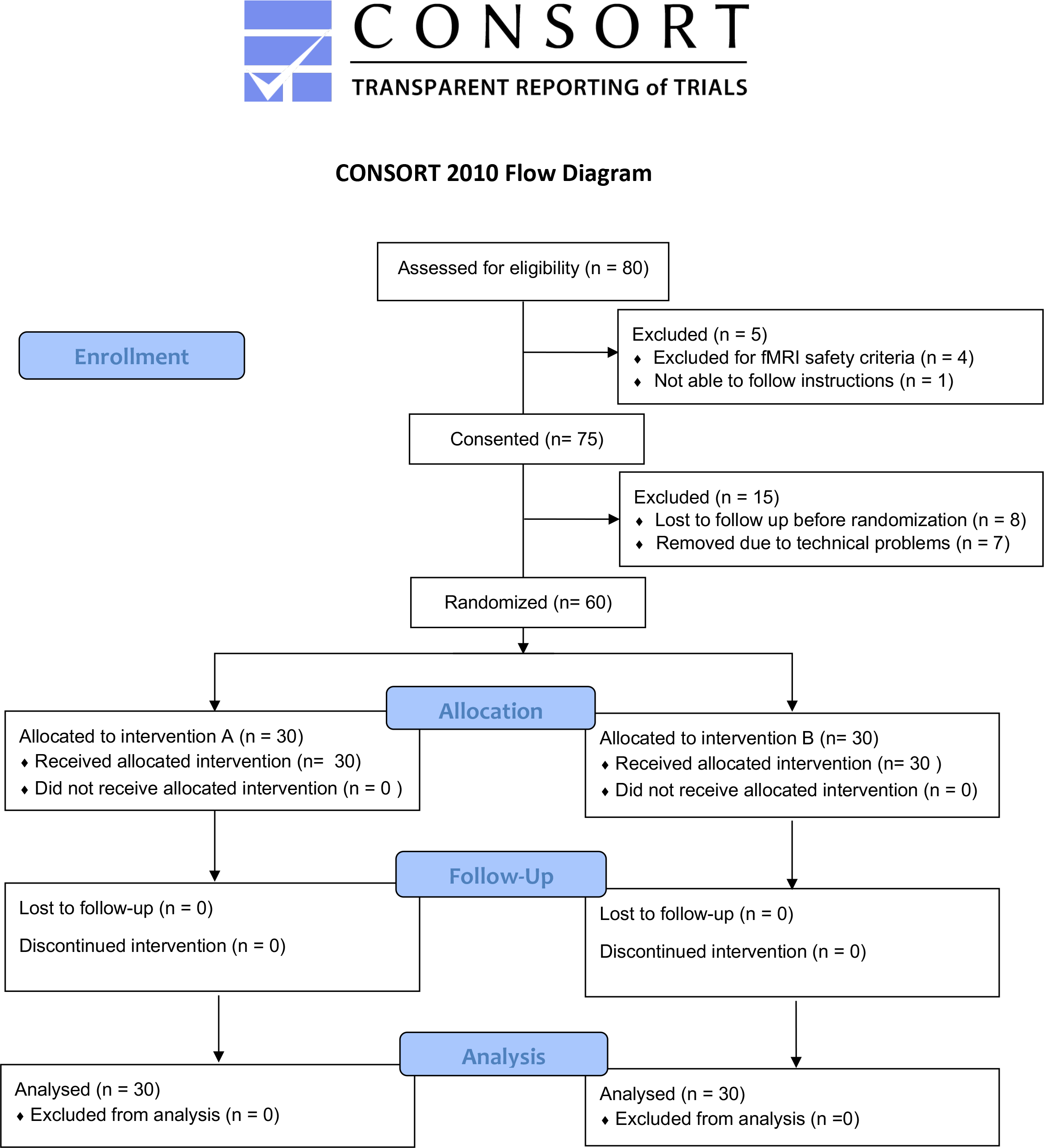

## Supplementary Materials

**Figure S1:**
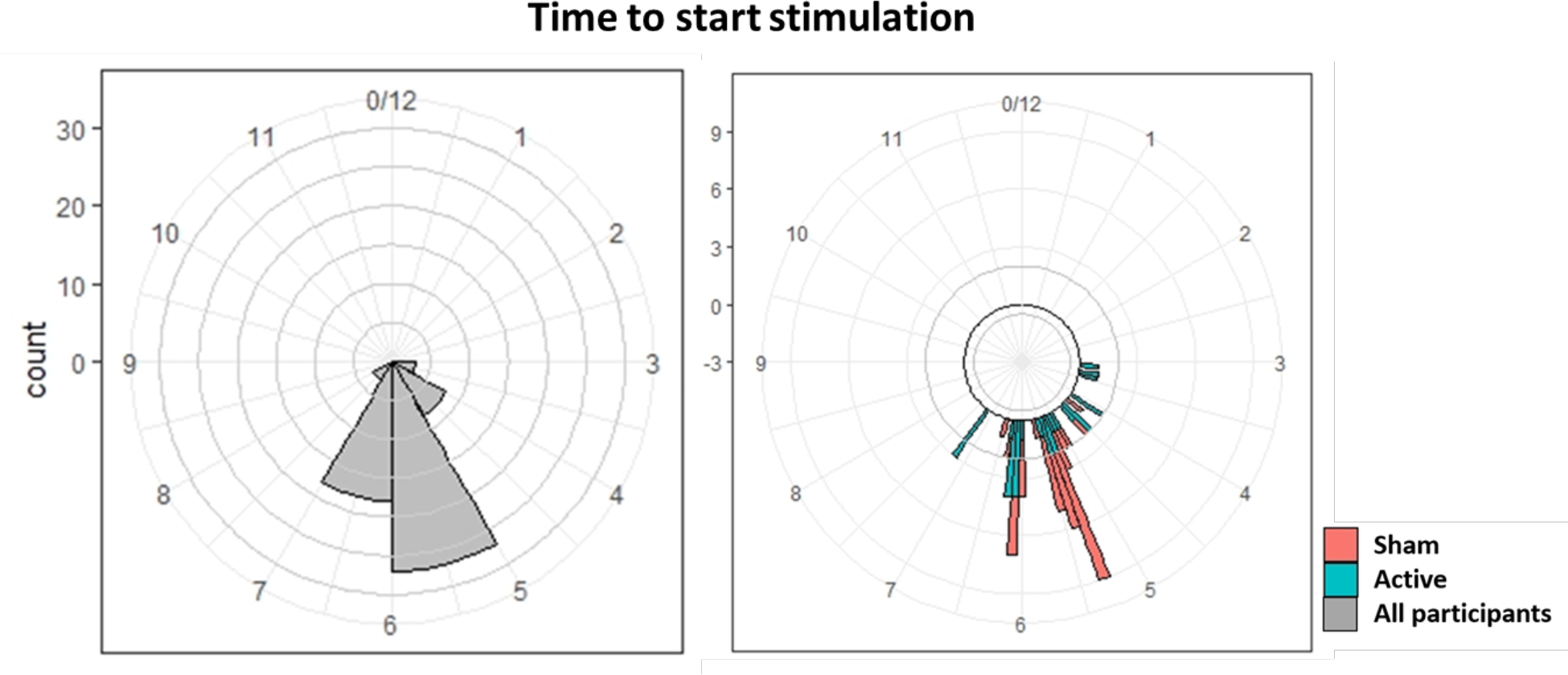
Distribution of the time of the day that stimulation was applied. Time to start of brain stimulation, as a critical factor that can affect stimulation outcomes and variation across the population, is visualized over a clock map for all participant (left side of figure) and each group separately (right side of figure). Each number in the figure is related to the time to start stimulation and circular histogram illustrating distribution of time throughout the day across all subjects. All participants received stimulation in the afternoon (from 3:00 pm to 7:30 pm).

**Figure S2.**
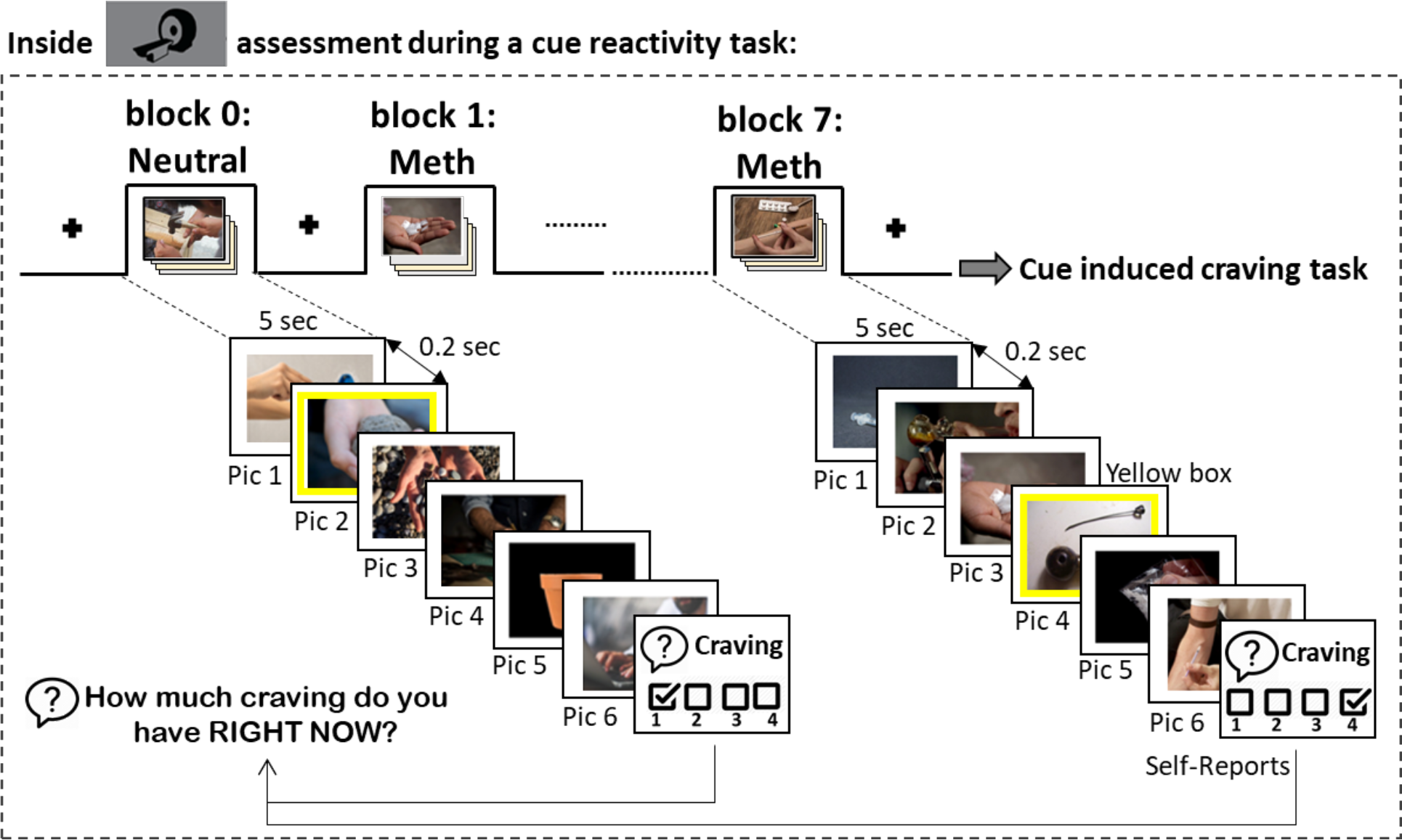
fMRI drug cue reactivity task paradigm. Schematic illustration of fMRI block design including examples of neutral stimuli and methamphetamine cues. Total task time was approximately 6 minutes consist of 4 neutral and 4 meth picture blocks. Each block incorporated a series of 6 pictures of the same category (meth or neutral) and were presented for 5 sec each with a 0.2 sec inter-stimulus interval. A visual fixation point was presented for 8 to 12 sec between each block. Each meth and neutral block were followed by meth craving inquiry in which participants were asked to rate their current meth craving level on a 1 to 4 rating scale (1 lowest to 4 highest). Participants’ response times to craving rating was also recorded. A yellow box was randomly presented around one of the six pictures in each block. Participants were asked to press a button as soon as they saw the box and their reaction time was recorded.

**Figure S3.**
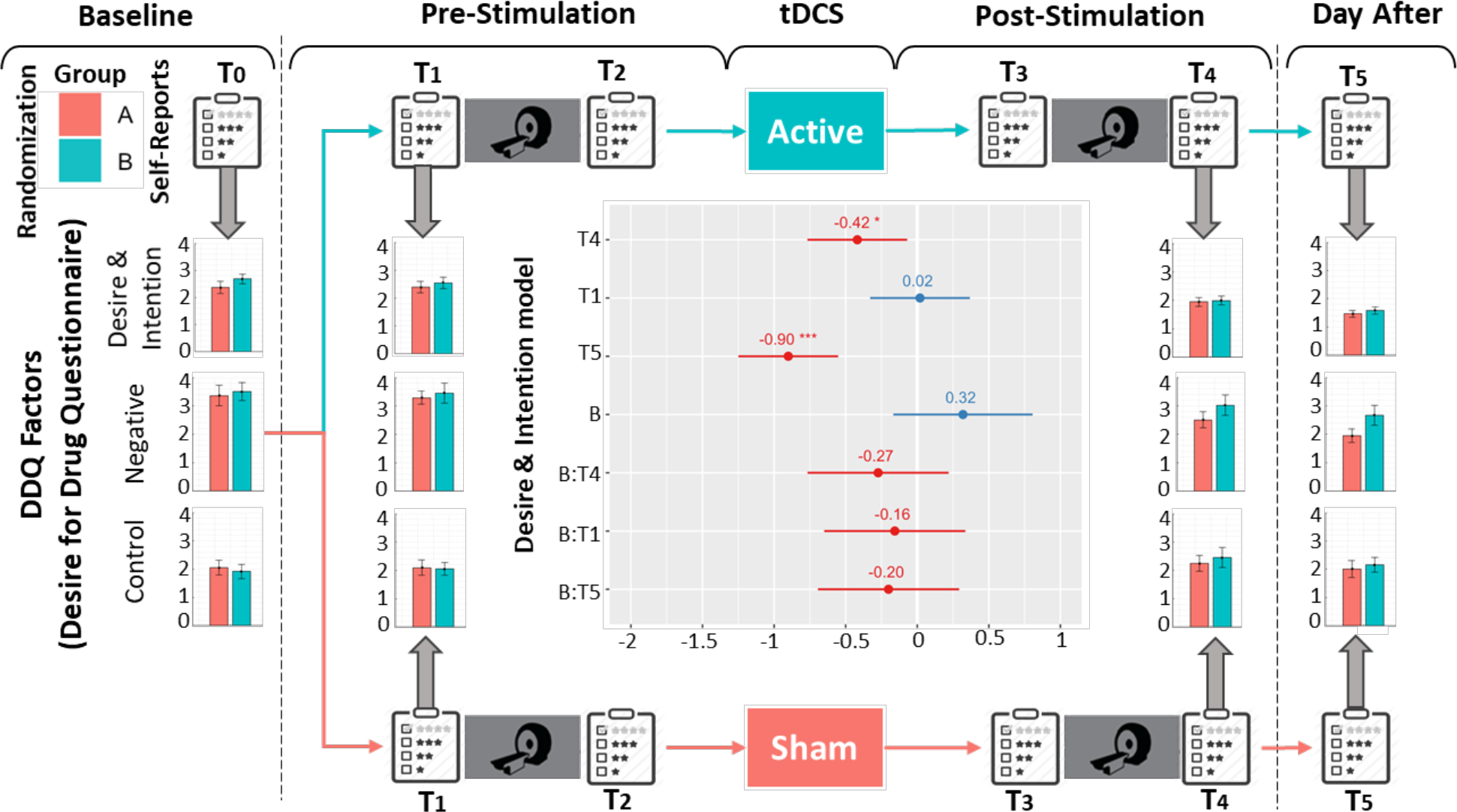
DDQ factors. Desire for Drug with three main factors including desire and intention (first row of bars), negative (second row of bars), and control (third row of bars) were collected at four time points: baseline (T0), pre stimulation before neuroimaging data collection (T1), post stimulation after neuroimaging data collection (T4), and the day after stimulation (T5). Linear mixed effect (LME) models were used with time, group, and time by group interaction as fixed effects and subjects as random effect. Significant (p uncorrected < 0.05) effect of time was found for all three factors. However, no effect of time or time by group interaction was found (results for desire and intention is represented in the centers of the figure with significant effect of T4 and T5, two other factors showed similar results).

**Figure S4:**
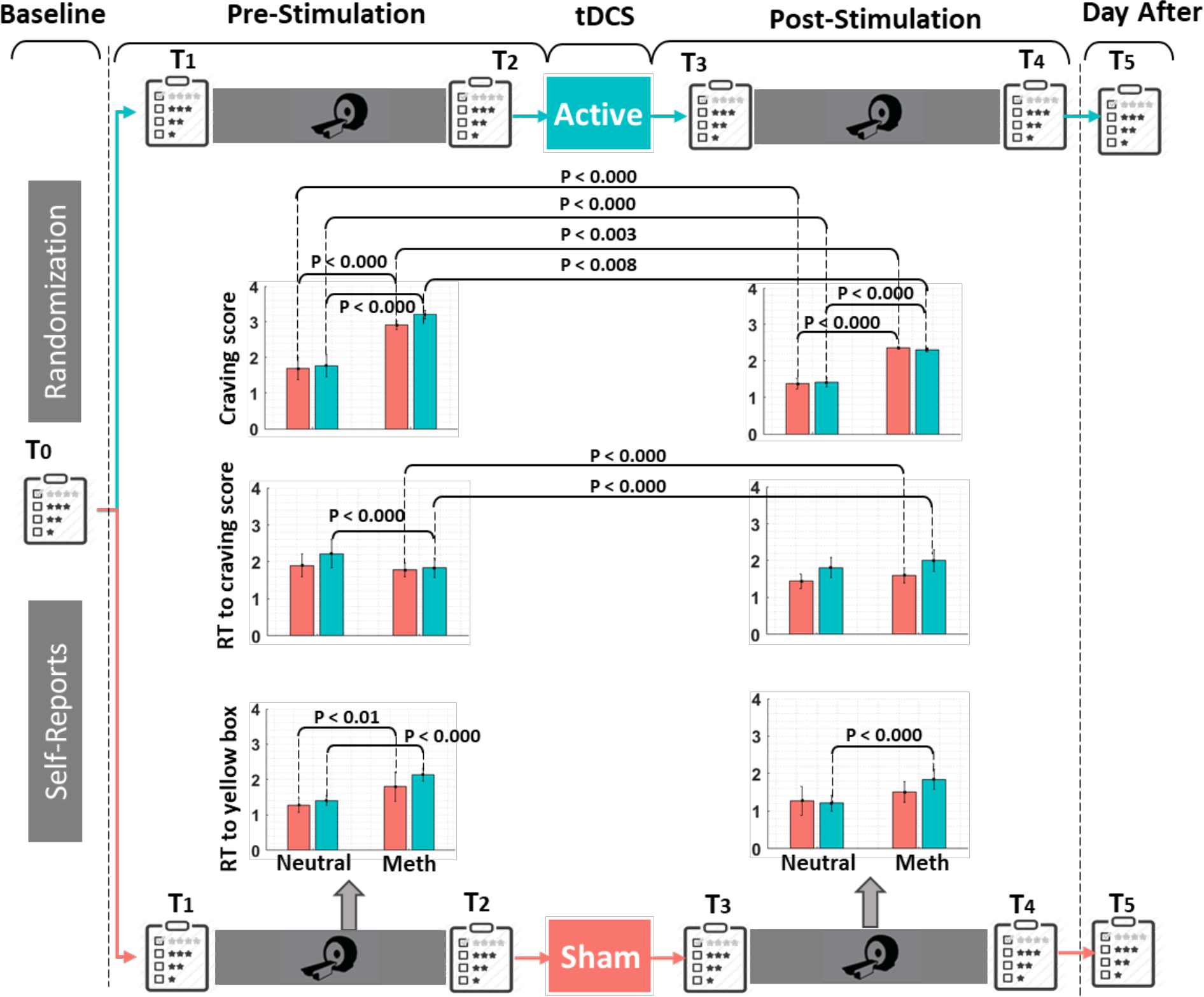
Inside scanner craving score and response time. Participants rated their craving levels during task-based fMRI. Response times to yellow boxes that were presented randomly around figures were recorded. Bar plots show averaged craving scores (first row), response time to craving score (second row), and reaction time to yellow boxes (third row) in each block (Meth or neutral) and error bars show standard error. Significant differences (p uncorrected < 0.05) are reported above bar charts.

**Figure S5:**
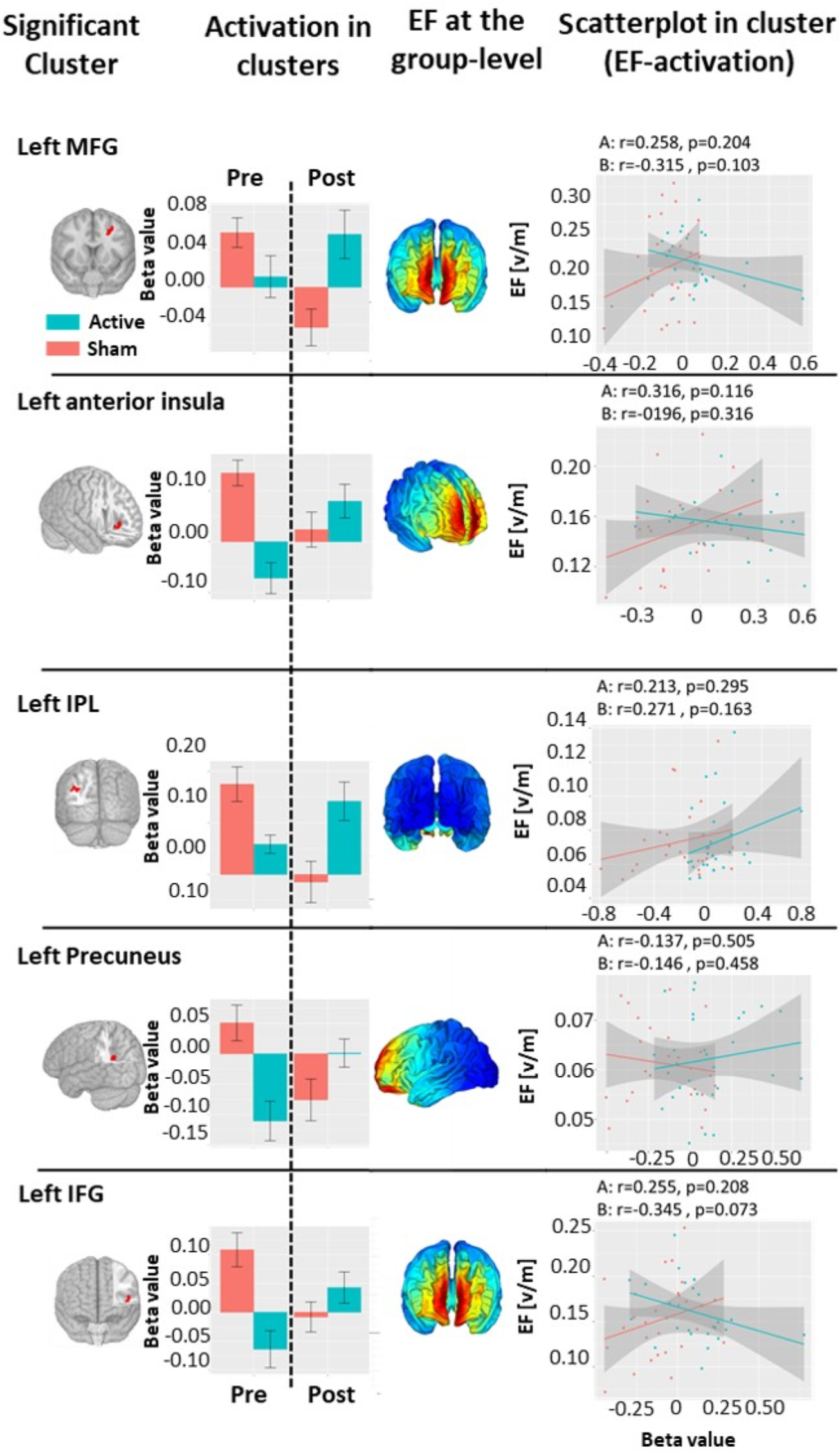
Activation and electric fields within clusters with significant time by group interaction. **<u>First column</u>**: clusters with significant (voxel level threshold: p uncorrected < 0.005, cluster size threshold > 40) time (pre vs post stimulation) by group (active vs sham) interaction visualized over the 3D MNI space. **<u>Second column:</u>** Bars show average beta values within clusters and error bars show standard error for (in blue) and sham (in red) group in pre (left side of the dashed line) and post (right side of the dashed line) neuroimaging data acquisition. **<u>Third column:</u>** electric field (EF) distribution patterns at the group-level in fsaverage space. **<u>Fourth column:</u>** Scatter plots for group-wise correlation between functional activity and averaged EF within the clusters. R and uncorrected p values are reported above each scatter plots. Regression models with two separate regressors (group and EF as regressors and changes in cluster activation as dependent variable) showed a significant effect of group in all 5 clusters.

**Figure S6:**
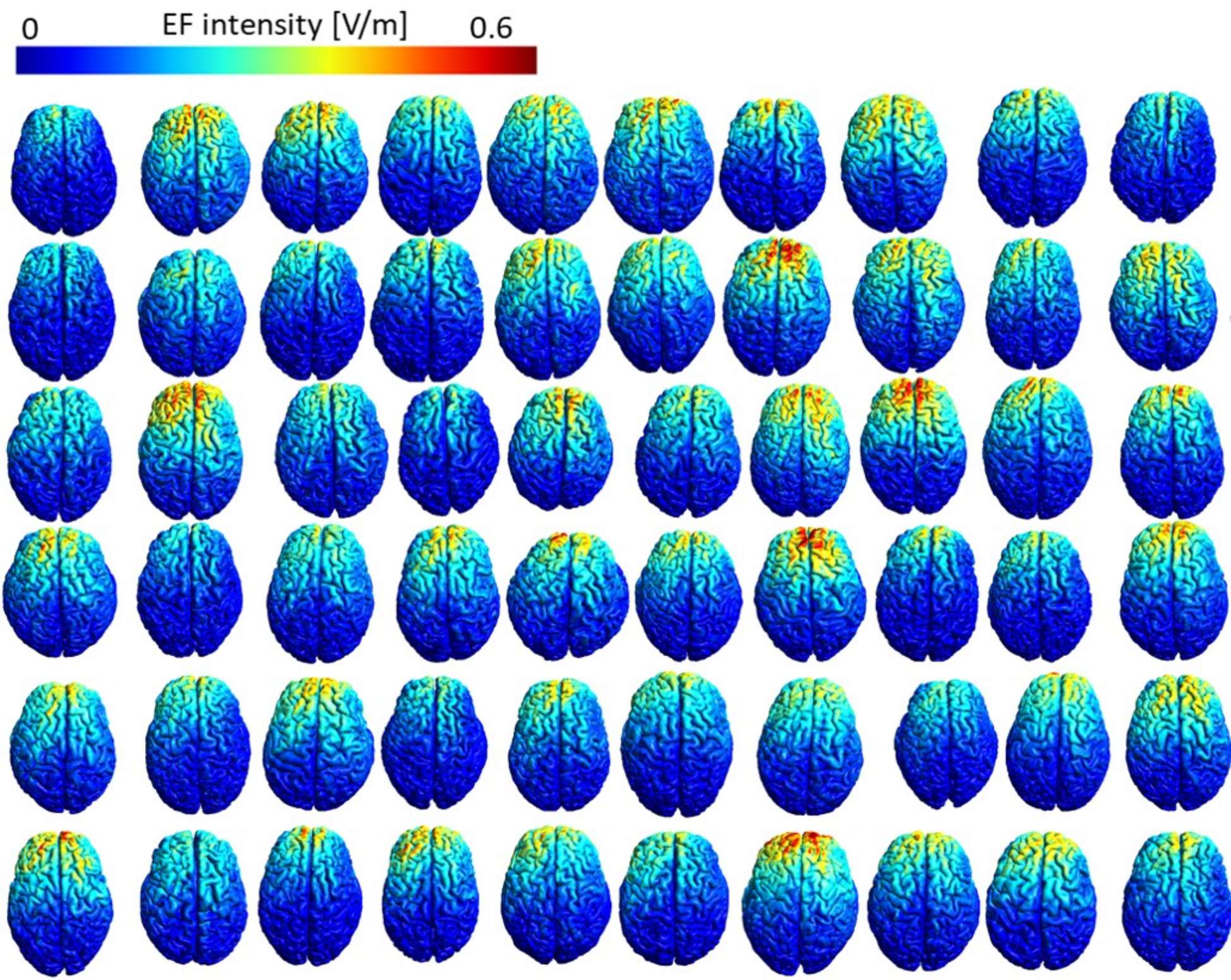
Individualized computational head models. Electric field (EF) distribution patterns of 60 participants for unilateral right DLPFC stimulation with anode/cathode over F4/Fp1 in individual native space with 2 mA current intensity. Abbreviation: EF: electric field, DLPFC: dorsolateral prefrontal cortex.

**Table S1:**
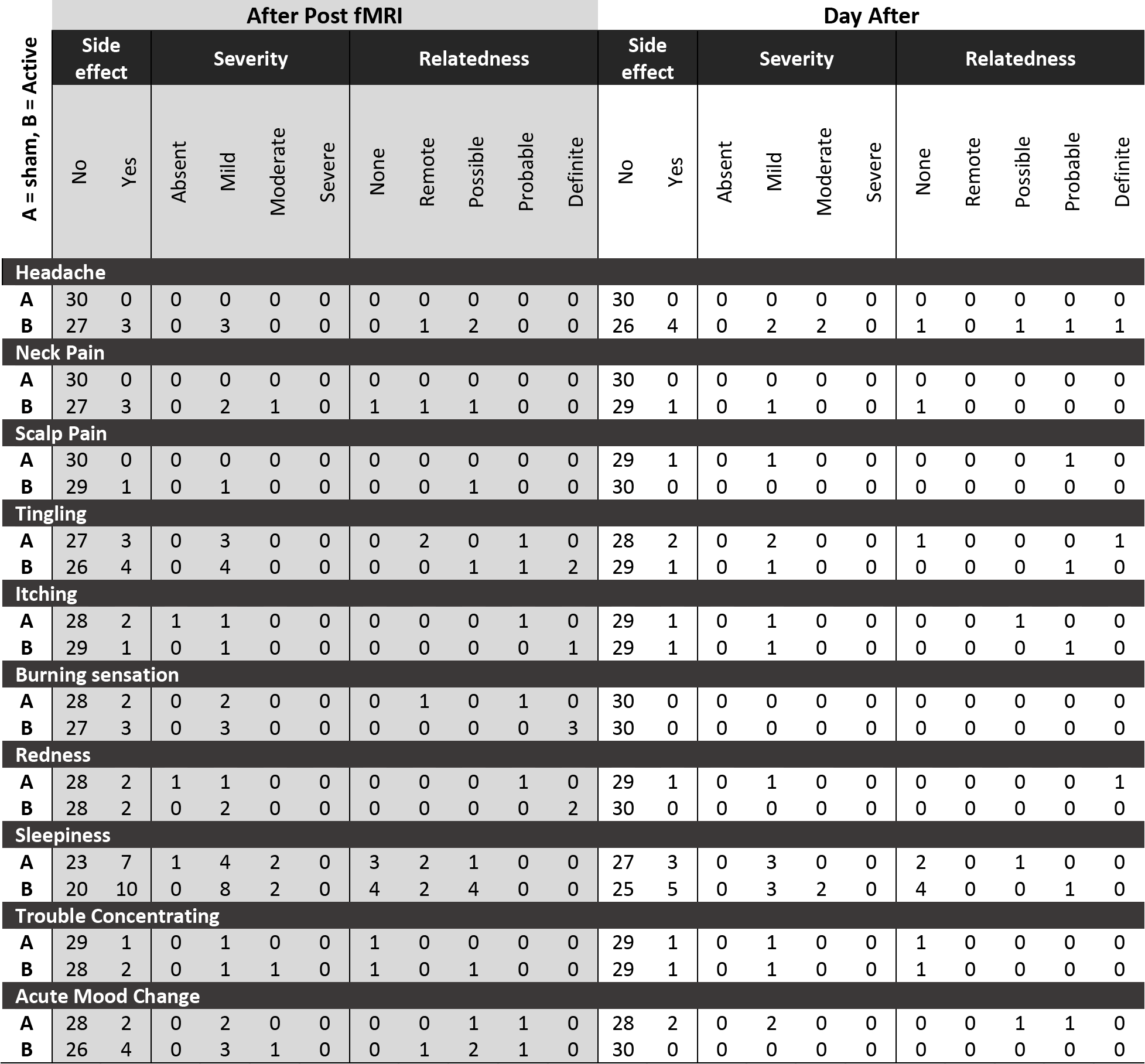
Side effect severity and perceived relatedness to the stimulation. After post-stimulation fMRI and the day after the stimulation participants were asked to determine if they had any side effects including headache, neck pain, scalp pain, tingling, itching, burning sensation, redness, sleepiness, trouble concentrating, and acute mood change by Yes or No (e.g., “Do you have a headache? Yes/No”). If they experienced any side effects after stimulation, they were asked to describe the severity (e.g., “Please describe the severity: absent/mild/moderate/severe”). Furthermore, they were asked if they thought the side effect was related to tDCS. Their answerers were quantified at five levels (e.g., “Is this related to tDCS? None/remote/possible/probable/definite”)

## References

Alireza Shahbabaie, Mehrshad Golesorkhi, Behnam Zamanian, Mitra Ebrahimpoor, Fatemeh Keshvari, Vahid Nejati, Felipe Fregni, Hamed Ekhtiari. 2014. “State dependent effect of transcranial direct current stimulation (tDCS) on methamphetamine craving.” International Journal of Neuropsychopharmacology 17 (10):1591–1598.

Bashir, Shahid, and Woo-Kyoung Yoo. 2016. “Neuromodulation for addiction by transcranial direct current stimulation: opportunities and challenges.” Annals of neurosciences 23 (4):241–245.

Batista, Edson Kruger, Jaisa Klauss, Felipe Fregni, Michael A Nitsche, and Ester Miyuki Nakamura-Palacios. 2015. “A randomized placebo-controlled trial of targeted prefrontal cortex modulation with bilateral tDCS in patients with crack-cocaine dependence.” International Journal of Neuropsychopharmacology 18 (12):pyv066.

Bikson, Marom, and Asif Rahman. 2013. “Origins of specificity during tDCS: anatomical, activity-selective, and input-bias mechanisms.” Frontiers in human neuroscience 7:688.

Boggio, Paulo S, Soroush Zaghi, Ana Beatriz Villani, Shirley Fecteau, Alvaro Pascual-Leone, and Felipe Fregni. 2010. “Modulation of risk-taking in marijuana users by transcranial direct current stimulation (tDCS) of the dorsolateral prefrontal cortex (DLPFC).” Drug and alcohol dependence 112 (3):220–225.

Boroda, Elias, Scott R Sponheim, Mark Fiecas, and Kelvin O Lim. 2020. “Transcranial direct current stimulation (tDCS) elicits stimulus-specific enhancement of cortical plasticity.” Neuroimage 211:116598.

Button, Katherine S, John PA Ioannidis, Claire Mokrysz, Brian A Nosek, Jonathan Flint, Emma SJ Robinson, and Marcus R Munafò. 2013. “Power failure: why small sample size undermines the reliability of neuroscience.” Nature reviews neuroscience 14 (5):365–376.

Cash, Robin FH, Luca Cocchi, Jinglei Lv, Paul B Fitzgerald, and Andrew Zalesky. 2020. “Functional Magnetic Resonance Imaging–Guided Personalization of Transcranial Magnetic Stimulation Treatment for Depression.” JAMA psychiatry.

Cleophas, Ton JM, and Ed M de Vogel. 1998. “Crossover studies are a better format for comparing equivalent treatments than parallel-group studies.” Pharmacy World and Science 20 (3):113–117.

Courtney, K. E., D. G. Ghahremani, and L. A. Ray. 2016. “The Effects of Pharmacological Opioid Blockade on Neural Measures of Drug Cue-Reactivity in Humans.” Neuropsychopharmacology 41 (12):2872–2881. doi: 10.1038/npp.2016.99.

Courtney, Kelly E, Joseph P Schacht, Kent Hutchison, Daniel JO Roche, and Lara A Ray. 2016. “Neural substrates of cue reactivity: association with treatment outcomes and relapse.” Addiction biology 21 (1):3–22.

Ekhtiari, H., P. Nasseri, F. Yavari, A. Mokri, and J. Monterosso. 2016. “Neuroscience of drug craving for addiction medicine: From circuits to therapies.” Prog Brain Res 223:115–41. doi: 10.1016/bs.pbr.2015.10.002.

Ekhtiari, Hamed, Rayus Kuplicki, Robin Aupperle, and Martin P Paulus. 2020. “It is Never as Good the Second Time Around: Brain Areas Involved in Salience Processing Habituate During Repeated Drug Cue Exposure in Methamphetamine and Opioid Users.” bioRxiv.

Ekhtiari, Hamed, Rayus Kuplicki, Asheema Pruthi, and Martin Paulus. 2020. “Methamphetamine and Opioid Cue Database (MOCD): development and validation.” Drug and alcohol dependence 209:107941.

Ekhtiari, Hamed, Hosna Tavakoli, Giovanni Addolorato, Chris Baeken, Antonello Bonci, Salvatore Campanella, Luis Castelo-Branco, Gaëlle Challet-Bouju, Vincent P Clark, and Eric Claus. 2019a. “Transcranial electrical and magnetic stimulation (tES and TMS) for addiction medicine: a consensus paper on the present state of the science and the road ahead.” Neuroscience & Biobehavioral Reviews 104:118–140.

Ekhtiari, Hamed, Hosna Tavakoli, Giovanni Addolorato, Chris Baeken, Antonello Bonci, Salvatore Campanella, Luis Castelo-Branco, Gaëlle Challet-Bouju, Vincent P Clark, and Eric Claus. 2019b. “Transcranial electrical and magnetic stimulation (tES and TMS) for addiction medicine: A consensus paper on the present state of the science and the road ahead.” Neuroscience & Biobehavioral Reviews.

Ekhtiari, Hamed, Mehran Zare-Bidoky, Arshiya Sangchooli, Amy C Janes, Marc J Kaufman, Jason Oliver, James J Prisciandaro, Torsten Wüstenberg, Raymond F Anton, and Patrick Bach. 2020. “A Methodological Checklist for fMRI Drug Cue Reactivity Studies: Development and Expert Consensus.” medRxiv.

Enck, Paul, Sibylle Klosterhalfen, and Stephan Zipfel. 2011. “Novel study designs to investigate the placebo response.” BMC medical research methodology 11 (1):1–8.

Esmaeilpour, Zeinab, A Duke Shereen, Peyman Ghobadi-Azari, Abhishek Datta, Adam J Woods, Maria Ironside, Jacinta O’Shea, Ulrich Kirk, Marom Bikson, and Hamed Ekhtiari. 2019. “Methodology for tDCS integration with fMRI.” medRxiv:19006288.

Fan, Lingzhong, Hai Li, Junjie Zhuo, Yu Zhang, Jiaojian Wang, Liangfu Chen, Zhengyi Yang, Congying Chu, Sangma Xie, and Angela R Laird. 2016. “The human brainnetome atlas: a new brain atlas based on connectional architecture.” Cerebral cortex 26 (8):3508–3526.

Fonteneau, Clara, Marine Mondino, Martijn Arns, Chris Baeken, Marom Bikson, Andre R Brunoni, Matthew J Burke, Tuomas Neuvonen, Frank Padberg, and Alvaro Pascual-Leone. 2019. “Sham tDCS: A hidden source of variability? Reflections for further blinded, controlled trials.” Brain stimulation 12 (3):668–673.

Franken, Ingmar HA, Vincent M Hendriks, and Wim van den Brink. 2002. “Initial validation of two opiate craving questionnaires: the Obsessive Compulsive Drug Use Scale and the Desires for Drug Questionnaire.” Addictive behaviors 27 (5):675–685.

Furubayashi, Toshiaki, Yasuo Terao, Noritoshi Arai, Shingo Okabe, Hitoshi Mochizuki, Ritsuko Hanajima, Masashi Hamada, Akihiro Yugeta, Satomi Inomata-Terada, and Yoshikazu Ugawa. 2008. “Short and long duration transcranial direct current stimulation (tDCS) over the human hand motor area.” Experimental brain research 185 (2):279–286.

Ganho-Ávila, Ana, Raquel Guiomar, Valério Daniela, Óscar F Gonçalves, and Jorge Almeida. 2019. “Cathodal tDCS impacts the fear network activity and connectivity patterns during a therapy-like extinction session.” L’Encéphale 45:S67.

Ghobadi-Azbari, Peyman, Asif Jamil, Fatemeh Yavari, Zeinab Esmaeilpour, Nastaran Malmir, Rasoul Mahdavifar-Khayati, Ghazaleh Soleimani, Yoon-Hee Cha, A Duke Shereen, and Michael A Nitsche. 2020. “fMRI and Transcranial Electrical Stimulation (tES): A systematic review of parameter space and outcomes.” Progress in Neuro-Psychopharmacology and Biological Psychiatry:110149.

Han, Beth, Jessica Cotto, Kathleen Etz, Emily B Einstein, Wilson M Compton, and Nora D Volkow. 2021. “Methamphetamine Overdose Deaths in the US by Sex and Race and Ethnicity.” JAMA psychiatry.

Hanlon, Colleen A, Logan T Dowdle, Nicole B Gibson, Xingbao Li, Sarah Hamilton, Melanie Canterberry, and Michaela Hoffman. 2018. “Cortical substrates of cue-reactivity in multiple substance dependent populations: transdiagnostic relevance of the medial prefrontal cortex.” Translational psychiatry 8 (1):1–8.

Jansen, Jochem M, Joost G Daams, Maarten WJ Koeter, Dick J Veltman, Wim van den Brink, and Anna E Goudriaan. 2013. “Effects of non-invasive neurostimulation on craving: a meta-analysis.” Neuroscience & Biobehavioral Reviews 37 (10):2472–2480.

Javadi, Amir Homayoun, Paul Cheng, and Vincent Walsh. 2012. “Short duration transcranial direct current stimulation (tDCS) modulates verbal memory.” Brain stimulation 5 (4):468–474.

Klauss, Jaisa, Quézia S Anders, Luna V Felippe, Michael A Nitsche, and Ester M Nakamura-Palacios. 2018. “Multiple sessions of transcranial direct current stimulation (tDCS) reduced craving and relapses for alcohol use: A randomized placebo-controlled trial in alcohol use disorder.” Frontiers in pharmacology 9:716.

Klauss, Jaisa, Leon Cleres Penido Pinheiro, Bruna Lima Silva Merlo, Gerson de Almeida Correia Santos, Felipe Fregni, Michael A Nitsche, and Ester Miyuki Nakamura-Palacios. 2014. “A randomized controlled trial of targeted prefrontal cortex modulation with tDCS in patients with alcohol dependence.” International Journal of Neuropsychopharmacology 17 (11):1793-1803.

Kroczek, AM, FB Häußinger, T Rohe, S Schneider, C Plewnia, A Batra, AJ Fallgatter, and A-C Ehlis. 2016. “Effects of transcranial direct current stimulation on craving, heart-rate variability and prefrontal hemodynamics during smoking cue exposure.” Drug and alcohol dependence 168:123–127.

Li, Lucia M, Ines R Violante, Rob Leech, Ewan Ross, Adam Hampshire, Alexander Opitz, John C Rothwell, David W Carmichael, and David J Sharp. 2019. “Brain state and polarity dependent modulation of brain networks by transcranial direct current stimulation.” Human brain mapping 40 (3):904–915.

Lupi, Matteo, Giovanni Martinotti, Rita Santacroce, Eduardo Cinosi, Maria Carlucci, Stefano Marini, Tiziano Acciavatti, and Massimo di Giannantonio. 2017. “Transcranial direct current stimulation in substance use disorders: a systematic review of scientific literature.” The Journal of ECT 33 (3):203–209.

Minarik, Tamas, Barbara Berger, Laura Althaus, Veronika Bader, Bianca Biebl, Franziska Brotzeller, Theodor Fusban, Jessica Hegemann, Lea Jesteadt, and Lukas Kalweit. 2016. “The importance of sample size for reproducibility of tDCS effects.” Frontiers in human neuroscience 10:453.

Moratalla, R, S Ares-Santos, and N Granado. 2014. “Neurotoxicity of methamphetamine.” Handbook of Neurotoxicity. New York: Springer:2207–30.

Neri, Francesco, Lucia Mencarelli, Arianna Menardi, Fabio Giovannelli, Simone Rossi, Giulia Sprugnoli, Alessandro Rossi, Alvaro Pascual-Leone, Ricardo Salvador, and Giulio Ruffini. 2020. “A novel tDCS sham approach based on model-driven controlled shunting.” Brain stimulation 13 (2):507–516.

Nikolin, Stevan, Donel Martin, Colleen K Loo, and Tjeerd W Boonstra. 2018. “Effects of TDCS dosage on working memory in healthy participants.” Brain stimulation 11 (3):518–527.

Rossini, Paolo Maria, D Burke, R Chen, LG Cohen, Zafiris Daskalakis, R Di Iorio, V Di Lazzaro, F Ferreri, PB Fitzgerald, and MS George. 2015. “Non-invasive electrical and magnetic stimulation of the brain, spinal cord, roots and peripheral nerves: basic principles and procedures for routine clinical and research application. An updated report from an IFCN Committee.” Clinical neurophysiology 126 (6):1071–1107.

Sale, Martin V, Michael C Ridding, and Michael A Nordstrom. 2007. “Factors influencing the magnitude and reproducibility of corticomotor excitability changes induced by paired associative stimulation.” Experimental brain research 181 (4):615–626.

Salehinejad, M, M Kuo, and M Nitsche. 2019. “The impact of chronotypes and time of the day on tDCS-induced motor cortex plasticity and cortical excitability.” *Brain Stimulation: Basic*, Translational, and Clinical Research in Neuromodulation 12 (2):421.

Sehatpour, Pejman, Clément Dondé, Matthew J Hoptman, Johanna Kreither, Devin Adair, Elisa Dias, Blair Vail, Stephanie Rohrig, Gail Silipo, and Javier Lopez-Calderon. 2020. “Network-level mechanisms underlying effects of transcranial direct current stimulation (tDCS) on visuomotor learning.” Neuroimage 223:117311.

Shaerzadeh, Fatemeh, Wolfgang J Streit, Soomaayeh Heysieattalab, and Habibeh Khoshbouei. 2018. “Methamphetamine neurotoxicity, microglia, and neuroinflammation.” Journal of neuroinflammation 15 (1):1–6.

Shahbabaie, Alireza, Mitra Ebrahimpoor, Ali Hariri, Michael A Nitsche, Javad Hatami, Emad Fatemizadeh, Mohammad Ali Oghabian, and Hamed Ekhtiari. 2018. “Transcranial DC stimulation modifies functional connectivity of large-scale brain networks in abstinent methamphetamine users.” Brain and behavior 8 (3):e00922.

Shahbabaie, Alireza, Mehrshad Golesorkhi, Behnam Zamanian, Mitra Ebrahimpoor, Fatemeh Keshvari, Vahid Nejati, Felipe Fregni, and Hamed Ekhtiari. 2014. “State dependent effect of transcranial direct current stimulation (tDCS) on methamphetamine craving.” International Journal of Neuropsychopharmacology 17 (10):1591–1598.

Sharifi-Fardshad, Mona, Mehdi Mehraban-Eshtehardi, Hassan Shams-Esfandabad, Schwann Shariatirad, Nader Molavi, and Peyman Hassani-Abharian. 2018. “Modulation of Drug Craving in Crystalline-Heroin Users by Transcranial Direct Current Stimulation of Dorsolateral Prefrontal Cortex.” Addiction & health 10 (3):173.

Smith, Robert C, Sylvia Boules, Sanela Mattiuz, Mary Youssef, Russell H Tobe, Henry Sershen, Abel Lajtha, Karen Nolan, Revital Amiaz, and John M Davis. 2015. “Effects of transcranial direct current stimulation (tDCS) on cognition, symptoms, and smoking in schizophrenia: a randomized controlled study.” Schizophrenia research 168 (1-2):260–266.

Supino, Phyllis G. 2012. “Fundamental issues in evaluating the impact of interventions: Sources and control of bias.” In Principles of research methodology, 79-110. Springer.

Trivedi, Madhukar H, Robrina Walker, Walter Ling, Adriane dela Cruz, Gaurav Sharma, Thomas Carmody, Udi E Ghitza, Aimee Wahle, Mora Kim, and Kathy Shores-Wilson. 2021. “Bupropion and Naltrexone in Methamphetamine Use Disorder.” New England Journal of Medicine 384 (2):140–153.

Verdejo-Garcia, Antonio, Valentina Lorenzetti, Victoria Manning, Hugh Piercy, Raimondo Bruno, Rob Hester, David Pennington, Serenella Tolomeo, Shalini Arunogiri, and Marsha E Bates. 2019. “A roadmap for integrating neuroscience into addiction treatment: a consensus of the neuroscience interest Group of the international society of addiction medicine.” Frontiers in psychiatry 10:877.

Whitfield-Gabrieli, Susan, and Alfonso Nieto-Castanon. 2012. “Conn: a functional connectivity toolbox for correlated and anticorrelated brain networks.” Brain connectivity 2 (3):125–141.

Wörsching, Jana, Frank Padberg, Konstantin Helbich, Alkomiet Hasan, Lena Koch, Stephan Goerigk, Sophia Stoecklein, Birgit Ertl-Wagner, and Daniel Keeser. 2017. “Test-retest reliability of prefrontal transcranial Direct Current Stimulation (tDCS) effects on functional MRI connectivity in healthy subjects.” Neuroimage 155:187–201.

Wu, Qiong, Chi-Fu Chang, Sisi Xi, I-Wen Huang, Zuxiang Liu, Chi-Hung Juan, Yanhong Wu, and Jin Fan. 2015. “A critical role of temporoparietal junction in the integration of top-down and bottom-up attentional control.” Human brain mapping 36 (11):4317–4333.

Xu, Jiansong, Felipe Fregni, Arthur L Brody, and Ardeshir S Rahman. 2013. “Transcranial direct current stimulation reduces negative affect but not cigarette craving in overnight abstinent smokers.” Frontiers in psychiatry 4:112.

Yang, Xue, Yong Wang, Qiyan Li, Yaxian Zhong, Liangpei Chen, Yajun Du, Jing He, Lvshuang Liao, Kun Xiong, and Chun-xia Yi. 2018. “The main molecular mechanisms underlying methamphetamine-induced neurotoxicity and implications for pharmacological treatment.” Frontiers in molecular neuroscience 11:186.

Zilverstand, Anna, Anna S Huang, Nelly Alia-Klein, and Rita Z Goldstein. 2018. “Neuroimaging impaired response inhibition and salience attribution in human drug addiction: a systematic review.” Neuron 98 (5):886–903.

